# Deciphering the Genetic Architecture of Parkinson’s Disease in India

**DOI:** 10.1101/2025.02.17.25322132

**Authors:** Asha Kishore, Ashwin Ashok Kumar Sreelatha, Amabel M. M. Tenghe, Rupam Borgohain, Divya Kalikavil Puthanveedu, Roopa Rajan, Madhusoodanan Urulangodi, Luis-Giraldo Gonzalez-Ricardo, Pramod Kumar Pal, Rukmini Mridula Kandadai, Saeideh Khodaee, Ravi Yadav, Sahil Mehta, Hrishikesh Kumar, Niraj Kumar, Prashanth Lingappa Kukkle, Soaham Dilip Desai, Kuldeep Shetty, Pettarusp Wadia, Annu Aggarwal, Pankaj Agarwal, Mirza Masoom Abbas, Gurusideshwar Mahadevappa Wali, Syam Krishnan, Divya Madathiparambil Radhakrishnan, Nitish Kamble, Achal Kumar Srivastava, Vivek Lal, Teresa Maria Costa Ferreira, Manas Chacko, Cibin Thoppil Raghavan, Gangadhara Sarma, Justin Solle, Brian Fiske, Amiya Thalakkatttu, Divyani Garg, Jens Krüger, Peter Lichtner, Dan Vitale, Mike Nalls, Cornelis Blauwendraat, Andy Singleton, Monojit Debnath, Swagata Sarkar, Sabbir Ansari, Sachin Adukia, Pravi Vidyadharan, R Kanthimathi, CKV Santhi, Tazeem Fathima Syed, Sindhuja Mohareer, Global Parkinson’s Genetics program (GP2), Lux-GIANT consortium, Manu Sharma

**Author notes:** Full list of the members in the supplementary document (Global Parkinson’s Genetics program (GP2)). Shared first authors (Asha Kishore, Ashwin Ashok Kumar Sreelatha, Amabel M. M. Tenghe). Corresponding authors 1. Manu Sharma, PhD, Head, Centre for Genetic Epidemiology, Institute for Clinical Epidemiology and Applied Biometry, University of Tübingen, Germany, 2. Asha Kishore MD, DM, FRCP, Director Parkinson and Movement Disorder Center, Aster Medcity, Kerala, India.

## Abstract

The genomic landscape of the Indian population, particularly for age-related disorders like Parkinson’s disease (PD) remains underrepresented in global research. Genetic variability in PD has been studied predominantly in European populations, offering limited insights into its role within the Indian population. To address this gap, we conducted the first pan-India genomic survey of PD involving 4,806 cases and 6,364 controls, complemented by a meta-analysis integrating summary statistics from a multi-ancestry PD meta-analysis (N=611,485). We further leveraged RNA-sequencing data from lymphoblastoid cell lines of 731 individuals from the 1000 Genomes project to evaluate the expression of key loci across global populations. Our findings reveal a higher genetic burden of PD in the Indian population compared to Europeans, accounting for ∼30% of the previously unexplained heritability. Thirteen genome-wide significant loci were identified, including two novel loci, with an additional three loci uncovered through meta-analysis. Polygenic risk score analysis showed moderate transferability from European populations. Our results highlight the importance of genetic loci in immune function, lipid metabolism and *SNCA* aggregation in PD pathogenesis, with gene expression variability emphasizing population-specific differences. We also established South Asia’s largest PD biobank, providing a foundation for patient-centric approaches to PD research and treatment in India.

## Introduction

With an estimated population of 1.48 billion, India ranks first in population density and is home to 17% of the global population. Despite this enormous demographic dividend that includes 4,500-5,000 anthropologically well-defined ethnolinguistic groups; the representation of genetic diversity in genomic studies has not matched the size and scale that India represents^1,2^. However, efforts are ongoing to offset these shortcomings and catalogue population-specific genetic variants to enhance the understanding of the myriad of chronic diseases in diverse populations^3–7^. To date, less than 1% of genome-wide association studies (GWAS) were conducted on the Indian population, and no study has yet represented the pan-Indian genetic architecture of neurodegenerative diseases, including sporadic PD^8^. Given the current global estimates, the 21st century will witness a considerable increase in PD, particularly in low-middle-income countries such as India^9^.

Genetic advancements in PD, including monogenic and sporadic forms, have primarily stemmed from studies conducted in European populations^8^. However, the transferability and the replicability of such findings in the Indian population remain untested. For example, while genes such as Parkin (*PRKN*) and to some extent *PINK1* have been implicated in familial PD, their role in the Indian population^10^ is limited, with other monogenic genes having a marginal impact^11^. Most studies in India have focussed on a candidate-gene approach for sporadic PD, often excluding the potential contribution of monogenic factors. Notably, a recent GWAS on the early onset of PD in India highlighted the role of *SNCA,* although the findings were based on a small sample size^12^. To strengthen these results, larger, well-powered cohorts are needed for validation^13^.

Genomics-driven research in India has been constrained by the lack of centralized infrastructure and sustainable funding mechanisms, limiting the ability to conduct comprehensive genomic surveys of PD in the Indian population. To address this critical gap the Luxembourg-German-Indian Alliance on Neurodegenerative Diseases and Therapeutics (Lux-GIANT; www.lux-giant.com) was established as a trilateral consortium. Supported by The Michael J. Fox Foundation for Parkinson’s Research and the Global Parkinson’s Genetics Program (www.gp2.org), we developed South Asia’s largest well-characterized PD biobank to promote and advance PD genetics research^14^. In brief, we implemented a hub-and-spoke model, establishing four major centralized nodes at leading tertiary referral hospital centres: Sree Chitra Tirunal Institute for Medical Sciences & Technology (SCTIMST, Thiruvananthapuram), All India Institute of Medical Sciences (AIIMS, Delhi), the National Institute of Mental Health and Neuro Sciences (NIMHANS, Bangalore), and Nizam’s Institute of Medical Sciences (NIMS, Hyderabad). These hubs are connected to 14 super-speciality hospitals functioning as sub-nodes, enabling comprehensive coverage of the PD population across India.

Leveraging this dedicated infrastructure, we report the largest PD GWAS conducted in India, consisting of 4,806 cases and 6,364 controls, ascertained and recruited over three years, from across the country. This study provides an unbiased survey of common risk variants for PD within the Indian population. We mapped common genetic associations with PD, identified novel putative loci, and confirmed the role of previously identified loci. In addition, we conducted cross-population polygenic prediction analyses comparing individuals of Indian and European ancestry. Finally, we provide an accessible framework that enables Indian investigators to use Lux-GIANT data to advance PD research locally and globally, addressing critical knowledge gaps and enhancing the representation of South Asian populations in global genomic studies.

## Results

### Genetic architecture of the Indian population

Two Indian clades representing the Indian population have been established in previous studies^15^. To minimize artefacts in our study, we investigated the relationship between the Indian population and global populations by integrating data from the Indian PD cohort with the 1000 Genomes Project (1000G) dataset and performing principal component analysis (PCA)^16,17^. Our data aligned with the South Asian reference population in the 1000G (**Figure 1A, 1B**). Given the considerable diversity of the Indian population, we further employed uniform manifold approximation and projection (UMAP), a non-linear dimensionality reduction approach to refine subpopulations and explore the fine-scale relationships between geography, genotypes, and phenotypes within the Indian population^18^. Using UMAP based on the top 10 principal components, we identified two major clusters within the Indian population, consistent with findings from previous studies^18^ (**Figure 1C**). The plots revealed that although two separate clusters were apparent, the distribution of our cohort from various regions of India was evenly spread. UMAP enabled finer resolution of population structure compared to PCA, uncovering clusters that might have been overlooked. This allowed us to minimize the impact of population stratification in downstream analyses.

**Figure 1:**
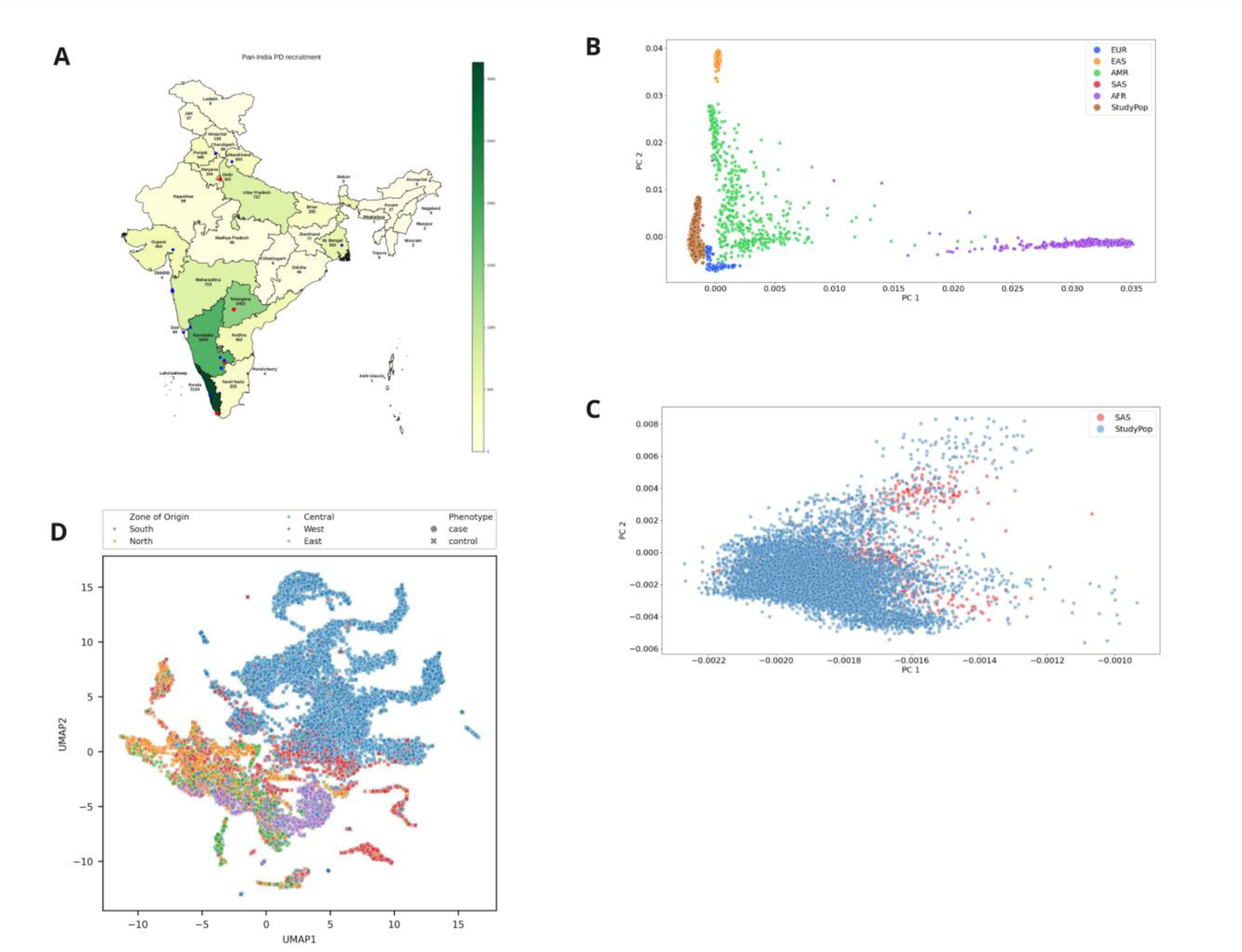
The distribution and genetic architecture of the PD cohort in India. (**A**) Pan-India recruitment of PD cases and age, gender-matched controls (**B**) PCA plot of the Indian population overlay with multi-ancestry samples from 1000 Genomes project. (**C**) PCA plot of Indian cohort with South Asian (SAS) super population of the 1000 Genomes project. The blue indicates the studied population, and the red indicates the reference population. Here, there is a complete overlap of the Indian PD cohort with the reference population. (**D**) UMAP plot computed with 10 principal components, min_dist=0.5 and n_neighbors=5. For reproducibility, a random seed was set equal to 42. There are clearly two different clades. Subject recruitment is evenly distributed, most of the recruitment is from the South because the major nodes were located South of India.

### PD GWAS in the Indian population

The final analysis included 4,806 cases and 6,364 controls after quality control (Methods). In this cohort, the mean age at onset for cases was 54.2 years (±11.87) while the mean age at recruitment for controls was 60.4 years (±15.28). The male-to-female ratio was 2:1, corresponding to 68% males and 32% females among cases, and 65% males and 35% females among controls.

Using a generalized linear model adjusted for age, sex, the top 10 PCs, and variants with minor allele frequency (MAF) > 1%, we identified 13 genome-wide significant loci in the Indian population (**Figure 2A**, **Table 1**). The genomic inflation factor (λ) was 1.095, and after applying genomic control (GC), λ_GC_ was reduced to 1.002 (**Supplementary Figure 1).** To ensure comparability with studies of similar scale, λ_GC_ was scaled to 1,000 cases and 1,000 controls, yielding 1.0002, indicating minimal inflation and robust adjustment for population structure.

**Figure 2:**
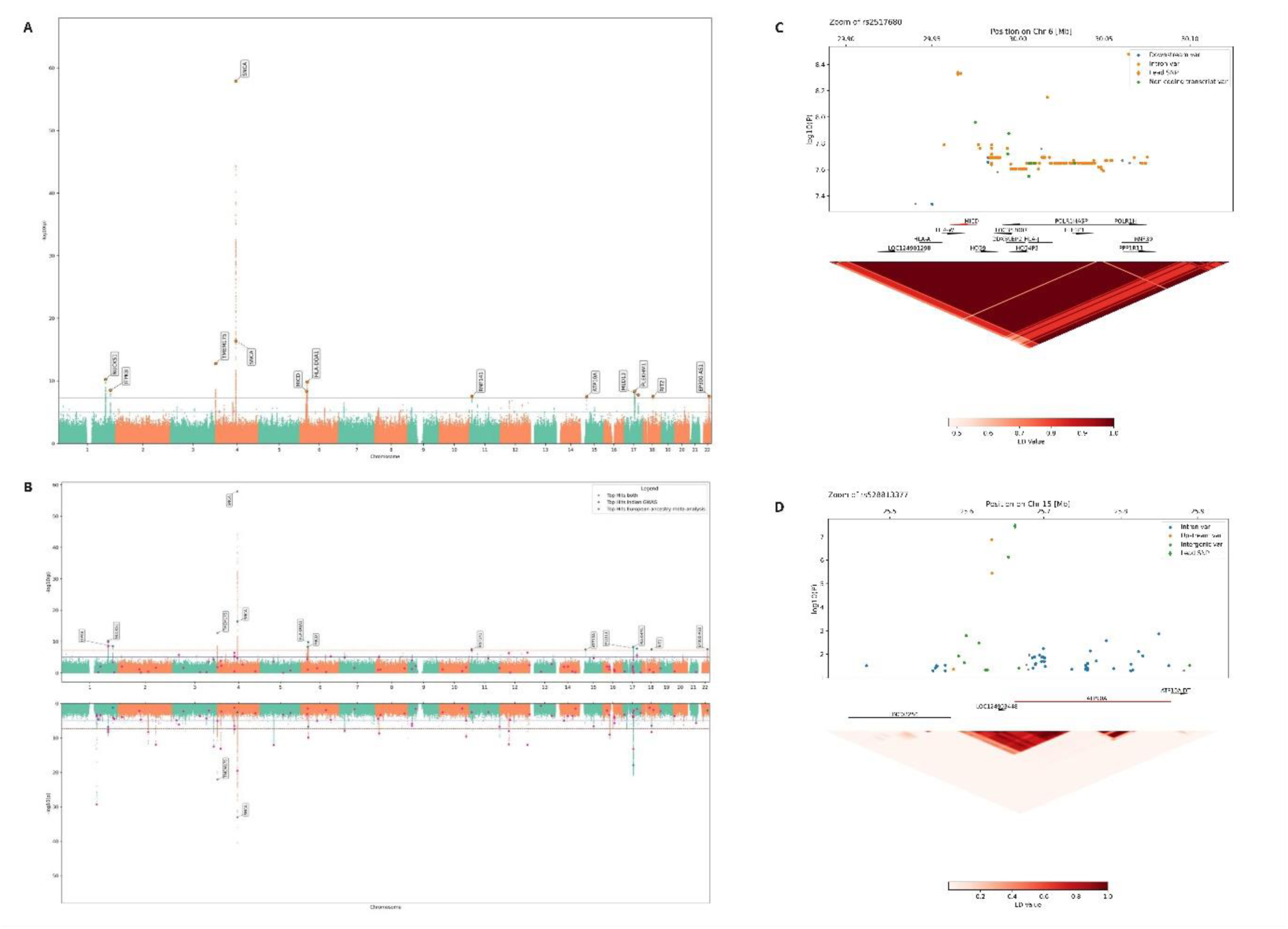
GWAS of Parkinson’s disease in the Indian population. (**A**) Manhattan plot of Indian GWAS where lead SNPs were annotated with the nearest gene. For annotation, the RefSeq update from 27.08.2024 was used. The X-axis indicates chromosomes and the Y-axis indicates –log(10) p-values. (**B**) Miami plot comparing Indian GWAS signals (upper panel) with European meta-analysis (lower panel). The X-axis indicates chromosomes, and the Y-axis –log (10) p-values. (**C - D**) Locus zoom and linkage disequilibrium plots of the two novel loci

**Table 1:**
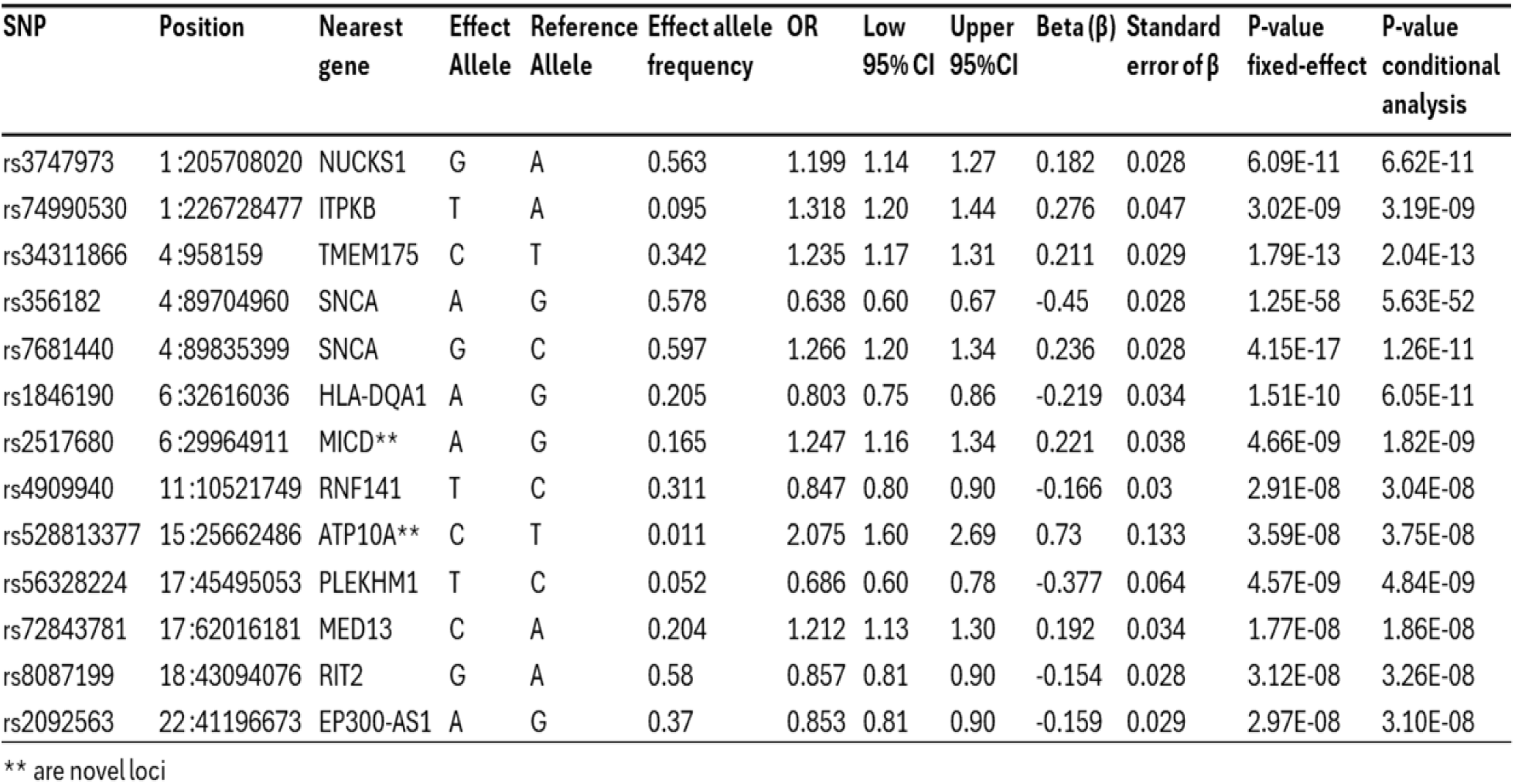
Genome-wide significant loci associated with Parkinson’s disease in the Indian population.

| SNP | Position | Nearest gene | Effect Allele | Reference Allele | Effect allele frequency | OR | Low 95% CI | Upper 95% CI | Beta ($\beta$) | Standard error of $\beta$ | P-value fixed-effect | P-value conditional analysis |
| --- | --- | --- | --- | --- | --- | --- | --- | --- | --- | --- | --- | --- |
| rs3747973 | 1:205708020 | NUCKS1 | G | A | 0.563 | 1.199 | 1.14 | 1.27 | 0.182 | 0.028 | 6.09E-11 | 6.62E-11 |
| rs74990530 | 1:226728477 | ITPKB | T | A | 0.095 | 1.318 | 1.20 | 1.44 | 0.276 | 0.047 | 3.02E-09 | 3.19E-09 |
| rs34311866 | 4:958159 | TMEM175 | C | T | 0.342 | 1.235 | 1.17 | 1.31 | 0.211 | 0.029 | 1.79E-13 | 2.04E-13 |
| rs356182 | 4:89704960 | SNCA | A | G | 0.578 | 0.638 | 0.60 | 0.67 | -0.45 | 0.028 | 1.25E-58 | 5.63E-52 |
| rs7681440 | 4:89835399 | SNCA | G | C | 0.597 | 1.266 | 1.20 | 1.34 | 0.236 | 0.028 | 4.15E-17 | 1.26E-11 |
| rs1846190 | 6:32616036 | HLA-DQA1 | A | G | 0.205 | 0.803 | 0.75 | 0.86 | -0.219 | 0.034 | 1.51E-10 | 6.05E-11 |
| rs2517680 | 6:29964911 | MICD** | A | G | 0.165 | 1.247 | 1.16 | 1.34 | 0.221 | 0.038 | 4.66E-09 | 1.82E-09 |
| rs4909940 | 11:10521749 | RNF141 | T | C | 0.311 | 0.847 | 0.80 | 0.90 | -0.166 | 0.03 | 2.91E-08 | 3.04E-08 |
| rs528813377 | 15:25662486 | ATP10A** | C | T | 0.011 | 2.075 | 1.60 | 2.69 | 0.73 | 0.133 | 3.59E-08 | 3.75E-08 |
| rs56328224 | 17:45495053 | PLEKHM1 | T | C | 0.052 | 0.686 | 0.60 | 0.78 | -0.377 | 0.064 | 4.57E-09 | 4.84E-09 |
| rs72843781 | 17:62016181 | MED13 | C | A | 0.204 | 1.212 | 1.13 | 1.30 | 0.192 | 0.034 | 1.77E-08 | 1.86E-08 |
| rs8087199 | 18:43094076 | RIT2 | G | A | 0.58 | 0.857 | 0.81 | 0.90 | -0.154 | 0.028 | 3.12E-08 | 3.26E-08 |
| rs2092563 | 22:41196673 | EP300-AS1 | A | G | 0.37 | 0.853 | 0.81 | 0.90 | -0.159 | 0.029 | 2.97E-08 | 3.10E-08 |

\*\* are novel loci

The effect allele frequency of the genome-wide significant loci ranges from 1% to 37% with effect estimates spanning from 0.85 to 2.08 in the Indian population (**Table 1**). Of the 13 identified loci, 2 are newly discovered (*ATP10A, and MICD)* and located more than 1Mb away from previously described variants^19^ suggesting they represent independent signals (Table 1). The four known loci (*NUCKS1, ITPKB, TMEM175, and RIT2*) exhibited contrasting effect size estimates compared to findings from a prior meta-analysis of European populations^19^ (**Table 1, Supplementary Figures 2-13**). Despite these differences in effect size, the variants exhibited consistent directionality with the results of the European meta-analysis. To further evaluate these observations, we compared the strength of genetic associations observed in the European meta-analyses with our Indian GWAS findings. Of the 90 SNPs reported in the European meta-analysis, 3 variants were absent, 5 variants had MAF < 1% and were excluded from further analyses, 6 variants were observed at genome-wide significance, 42 reached a significant threshold of *p* < 10^-5^ and 34 were observed at a nominal significance of *p* < 0.05 (**Figure 2B**). A total of 94.44% of SNPs exhibited the same directionality of effect estimates while 5.56% showed opposing directionality (**Supplementary Figure 14**). Power calculation indicated that our study had ∼95% power to detect the observed effect sizes (**Figure 3**).

**Figure 3:**
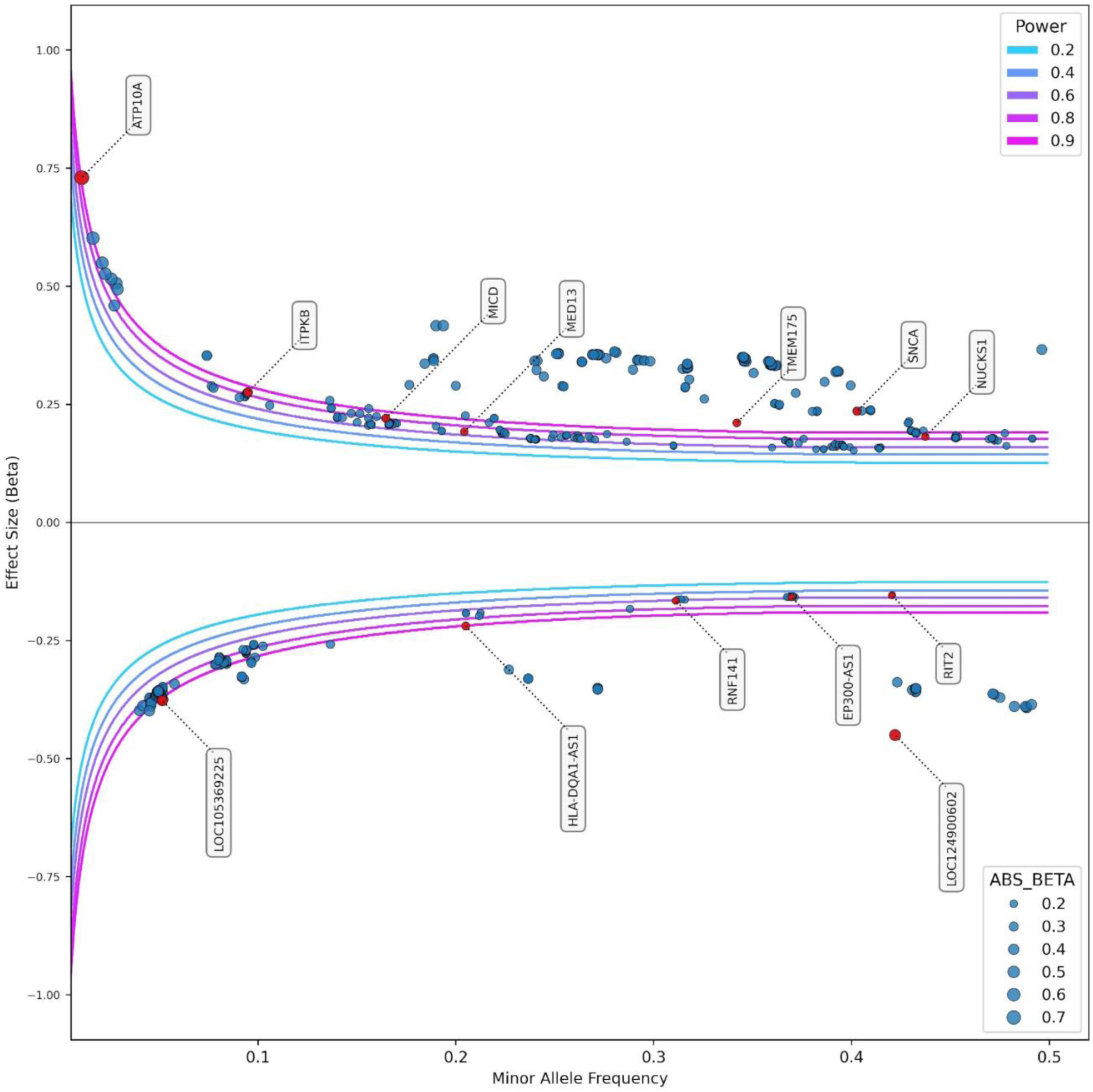
Power estimation under varied minor allele frequencies and effect estimates threshold for GWAS. The trumpet plot shows power estimates whereby minor allele frequencies are plotted against the effect estimates observed in our study. The different colour-coded lines indicate the power to detect the observed effect estimates. The X-axis represents the minor allele frequencies, and the Y-axis represents the effect estimates.

### The role of novel PD loci, LRRK2, GBA1, and MAPT in the Indian population

Of the two novel variants (ATP10A and MICD) identified, neither were reported in the European meta-analysis^19^ and one variant was non-significant in the multi-ancestry meta-analysis^20^. The directionality of effect estimates for these variants aligns with variants observed in a multi-ancestry meta-analysis, underscoring their relevance to PD. Similarly, two variants with the same directionality were observed in our study as in the previously published Indian GWAS, further emphasising their potential role in the Indian population^12^.

The global diversity array (GDA) designed to create a comprehensive global map of genetic variants, capturing ethnic diversity and enriched for neurodegenerative disease markers, referred to as GDA-NeuroBooster, was used to process our cohort^21^. Despite the overlap of known risk loci between the Indian and European populations, we did not observe the well-known bona fide loci (*LRRK2, GBA1* and *MAPT*) identified in the European population at a genome-wide level in the Indian population. Leveraging GDA-NeuroBooster allowed a systematic screening of our cohort for the known mutations and common risk variants at these loci.

The largest screening of the G2019S mutation excluded its role, as the G2019 mutation was absent in our cohort (4,806 cases and 6,364 controls). However, using a liberal significance threshold of (*P* < 1 × 10^−6^), we observed a clear signal at the *LRRK2* and *MAPT* loci, underscoring the potential contributions of common risk variants in the Indian population. Notably, the signal at the *LRRK2* locus in the Indian population is independent of the G2019S mutation, which is the major driver of PD in European populations^20,22^. A previous meta-analysis demonstrated that much of the *LRRK2* effect is driven by the G2019S mutation and the variant, rs76904798^19^. In contrast, the variant rs11175666, identified in the Indian population (*P* = 1.64 × 10⁻⁷), is located approximately 14Kb from rs76904798 (*P* = 5.013 × 10⁻^7^) and exhibits moderate linkage disequilibrium with it (r2 = 0.57, D*′* = 0.96). This finding suggests that the involvement of *LRRK2 in* the Indian population operates through a mechanism distinct from the G2019S-driven mechanism observed in European populations (**Supplementary Table 1**). Furthermore, the analysis of Asian-specific LRRK2 risk variants (G2385R and R1628P) identified six carriers of G2385R in our cohort, with the variant absent among 6,364 controls, indicating its extreme rarity in the Indian population. Similarly, R1628P was identified in five cases and one control. Despite the exceedingly low frequency of known Asian-specific *LRRK2* variants in the Indian population, the rare risk variants associated with *LRRK2*-dependent PD showed notable parallels with findings in other populations (**Supplementary Table 1**).

Our study also excluded the role of known *GBA1* mutations in the Indian population. However, p.E326K and p.T369M were associated with an increased risk for PD with *P* = 2.16 × 10⁻⁴ for p.E326K (OR=2.83, CI =1.53-5.22) and *P* = 8.80 × 10⁻⁴ for p.T369M (OR= 2.96, CI= 1.66-5.28). Both variants had an MAF of 0.0021 in the Indian population. The p.E326K variant was observed in 47 individuals (32 cases, 15 controls), while p.T369M was found in 50 individuals (34 cases, 16 controls). Notably, the effect estimates for both variants were higher than those reported in a large meta-analysis, underscoring their potential relevance in the Indian population^23^ (**Supplementary Table 1**).

The signal observed at the *MAPT* locus was borderline genome-wide significant (*P* = 5 × 10⁻⁸). The variant rs8070723, which tags the H1/H2 haplotype, revealed that the H2 haplotype is protective. Compared to the European population, the H2 haplotype occurs at a frequency of 4% in the Indian population and showed a robust association with reduced PD risk (OR = 0.73, CI = 0.63–0.84). In addition, we applied a six-variant SNP panel (rs1467967-rs242557-rs3785883-rs2471738-rs8070723-rs7521) to define MAPT H1 sub haplotypes^24^. While the H1-clade haplotypes occurred at varying frequencies, none were associated with PD in the Indian population (**Supplementary Tables 2-4**).

### Replication of Asian-specific loci in Indian PD population

The largest East Asian meta-analyses identified two novel Asian-specific loci for PD, *SV2C* (rs246814) and *WBSCR17 (rs9638616)*^25^. Similarly Chinese and South Korean GWAS identified the *HEATR6* gene and rs34778348 (LRRK2 G2385R) as associated with PD in their respective populations^26,27^. In our GWAS and meta-analysis, we did not observe association signals for *SV2C* or *WBSCR17*, suggesting that these loci are not involved in the Indian population. A recently published GWAS identified *CCDC85A* (rs59330234) as a novel locus in the Indian population^12^. However, this locus was found to be non-significant (*P* = 4.75E-01, SE = 0.0340), excluding the role of this gene in the Indian population. Furthermore, the same study identified the *BSN* gene as a rare gene associated with PD in the Indian population^12^. Upon examining a single marker within the *BSN* gene region, we did not observe an association with PD. Similarly, *HEATR6* was not associated with PD in our cohort^26^. Our comprehensive assessment of Asian-specific loci collectively excludes the involvement of these previously described loci in the Indian population (**Supplementary Table 5**).

### PD heritability in the Indian population

Our analysis revealed that 30% (95% CI= 26.37% - 33.85%) of the phenotypic variance observed in all PD cases can be attributed to genetic factors. For early-onset PD, the heritability was estimated at 46.8% (95% CI= 30.06%-63.6%) while for late-onset PD, it was 19% (95% CI= −0.89%-39.6%). Our findings for the first time suggest a significant genetic burden in the Indian population^19,28^. However, when focusing on the top SNPs, we did not observe a substantial increase in genetic variance (**Supplementary Table 6**).

### Meta-analysis of Indian and multi-ancestry PD GWAS

We performed a meta-analysis by incorporating previously published summary statistics from multi-ancestry PD GWAS^20^. When combined with our current study, we successfully replicated the two newly identified loci, reinforcing their universal role in PD pathogenesis. Notably, the conditional analysis uncovered 29 additional independent loci, 26 of which have been previously reported. Using a random effects model that accounts for heterogeneity, we identified three additional novel loci (*RLP8L1*, *TEC* and *DSCAM*) in the meta-analysis (**Supplementary Figure 15, Supplementary Table 10**).

### Polygenic risk score and PD

We evaluated the extent to which polygenic risk scores (PRS) derived from previously published GWAS summary statistics for European (EUR) and Indian (IND) cohorts could explain variation in PD risk^19,20^. Using the IND summary statistics as a base set, the best performing PRS model (*R²* = 0.88, p-value threshold (*P_T_*) < 5 × 10⁻⁸), included only three SNPs, explaining just 1% of PD risk (**Figure 4A**). To improve the prediction model, we applied a less stringent p-value threshold (*P*_T_ < 5 x 10^-6^), which increased the number of SNPs to 13 (**Supplementary Table 7A**). However, despite this inclusion of additional SNPs, the PRS risk variability showed no improvement (*R*^2^=0.84), suggesting that the small sample size in the base data may have limited the accuracy of risk prediction.

**Figure 4:**
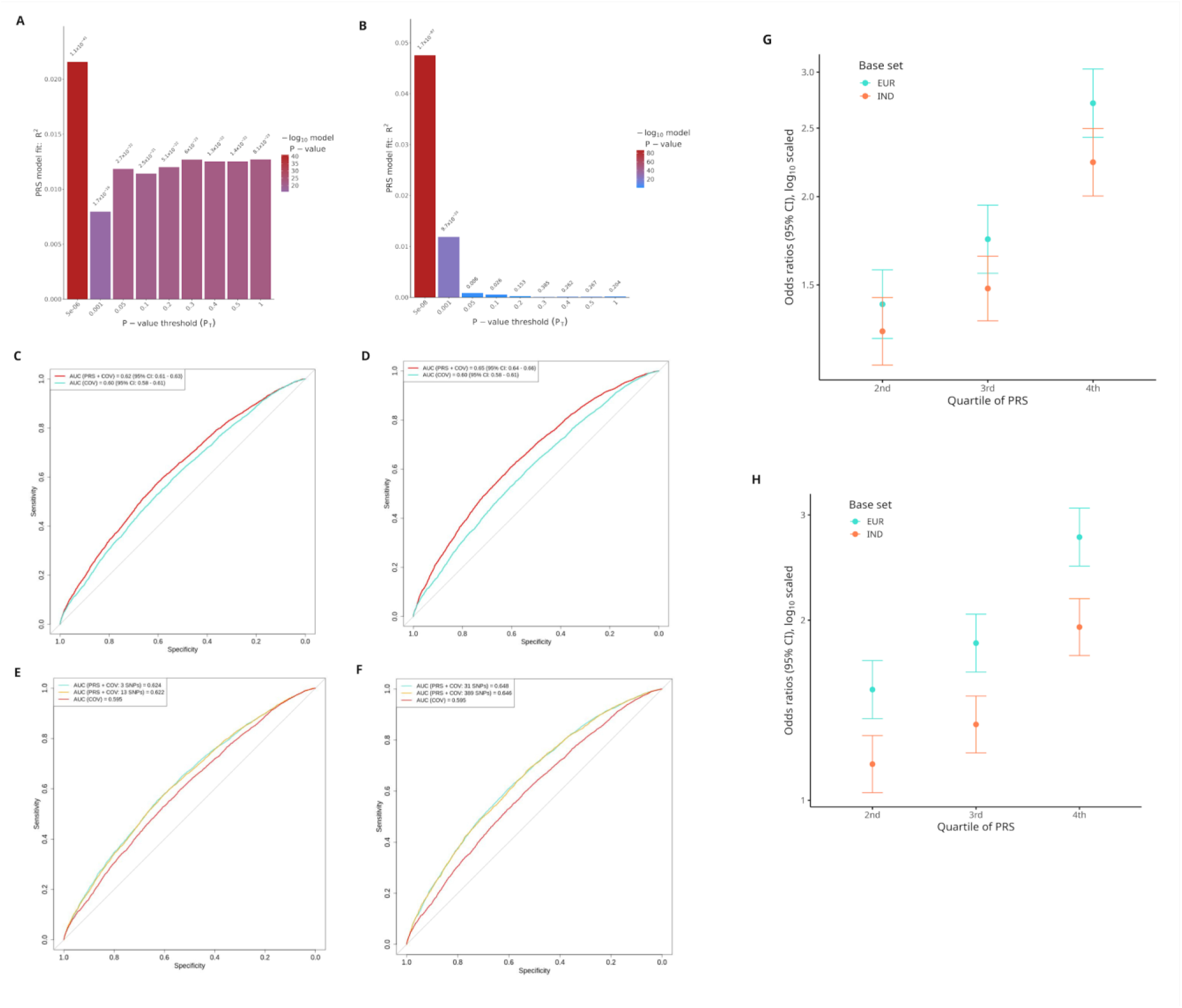
PRS model for comparing the transferability of PD risk between the Indian and the European populations. Polygenic risk score (PRS) prediction for Indian individuals, using European (EUR) or Indian (IND) base data. PRS was computed with GWAS summary statistics from EUR and IND populations as base sets. (**A**) is a bar plot of the R2 for the PRS models of different p-value thresholds evaluated in the IND base, and (**B**) is the bar plot for the EUR base? (**C**) Receiver-operator curves (ROC) for best-performing PRS were computed with 3 SNPs from the IND base. (**D**) ROC with 31 SNPs from the EUR base. (**E**) ROC for PRS from 3 and 13 IND base SNPs. (**F**) ROC for PRS from 31 and 389 EUR base SNPs. (**G**) is the odds ratio of developing PD for each quantile of PRS compared with the lowest quartile of genetic risk using 31 EUR base SNPs and 3 IND base SNPs, (**H**) odds ratio using 389 EUR base SNPs and 13 IND base SNPs.

In contrast, using the EUR base set increased the number of SNPs to 31, resulting in a marginal improvement of approximately 2% of PD risk prediction (*R*^2^=1.89, *P*_T_ < 5 x 10^-8^) (**Figure 4B**). This improvement is likely attributed to the larger sample size used to generate the EUR summary statistics. Similar to the IND base set, applying a lower p-value threshold (*P*_T_ < 5 x 10^-5^) further increased the number of SNPs to 389, but this adjustment did not enhance the predictive power for PD risk (**Supplementary Table 7A**).

Furthermore, the area under the curve (AUC) was comparable across both base sets. For the IND base set, the AUC was 62.4% for the 3 SNPs and 62.2% for the 13 SNPs. For the EUR base set, the AUC was 64.8% for the 31 SNPs and 64.6% for the 389 SNPs (**Figure 4C–F, Supplementary Table 7D**). Notably, risk stratification using PRS derived from the IND and EUR base sets revealed that individuals in the highest quartile had a PD odds ratio of 1.95 for the IND base set and 2.71 for the EUR base set, respectively (**Figure 4G-H, Supplementary Table 7 B-C**).

### Indian PD risk loci and gene expression in global populations

Using the MAGE dataset^29^, we evaluated the global expression of the top loci identified in our study across diverse populations (**Figure 5A, Supplementary Table 8A**). This analysis revealed significant expression differences for the top loci observed in our study across global populations^29^. *ITPKB* showed expression differences between South Asian (SAS) and multiple populations, including European (EUR), American (AMR) and African (AFR), as well as EUR and East Asian (EAS) populations. SNCA exhibited notable variation in expression between EUR populations and those from SAS, EAS and AFR. *HLA-DQA1* exhibited distinctive expression differences, particularly between SAS and AFR, with additional variability across AMR compared to EAS and AFR. *ATP10A* demonstrated expression differences between AFR and other populations, as well as between AMR with EAS and SAS.

**Figure 5:**
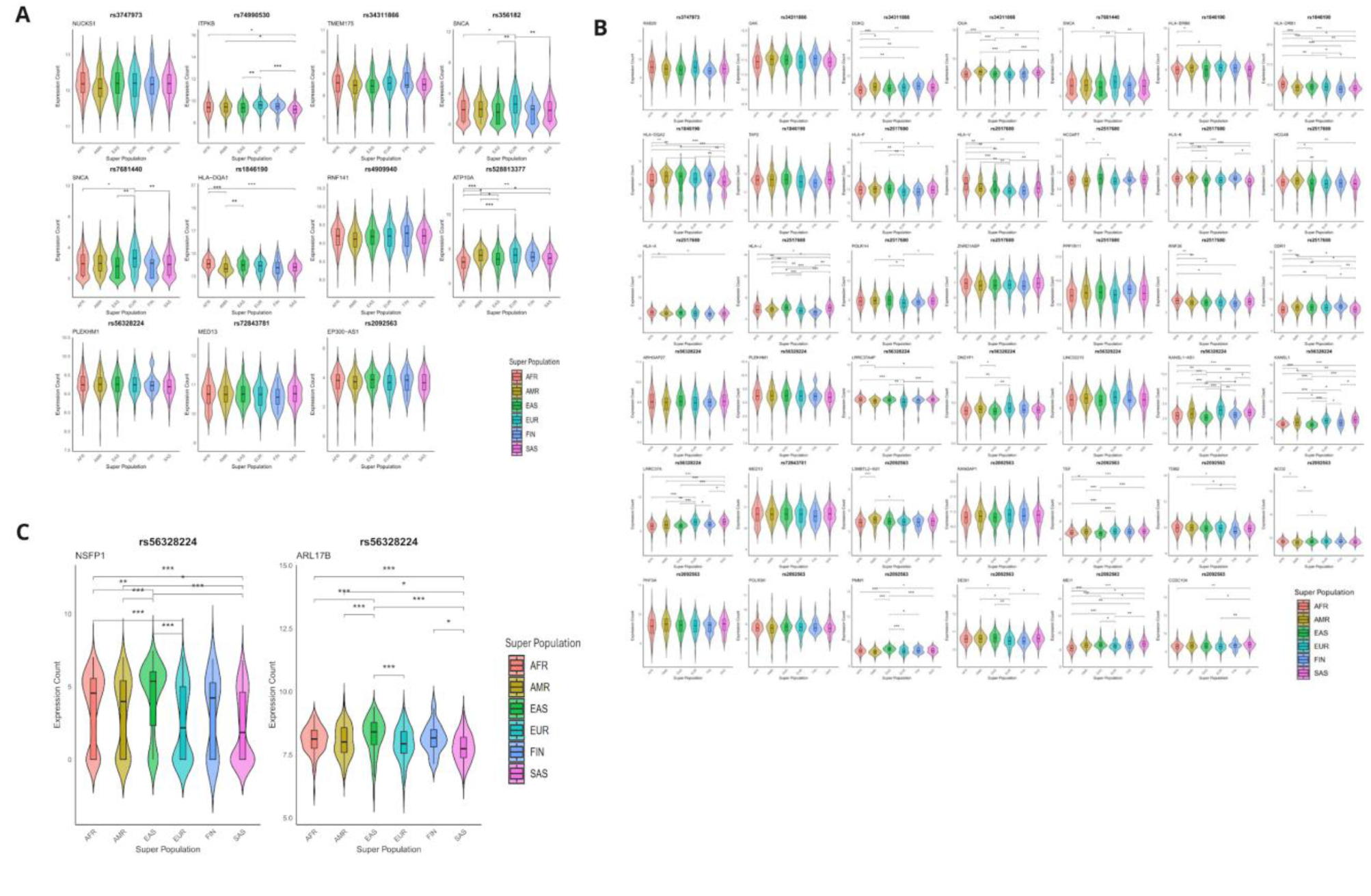
Assessment of gene expression and eQTL of genome-wide significant loci in the global populations. (**A**) Nearby genes associated with these loci. (**B**) Genes significantly linked to these loci in eQTL analysis (FastQTL). (**C**) Genes influenced by causal variant SNP rs56328224, identified via the SuSiE approach. *adjusted p-values < 0.05, **adjusted p-values < 0.01, ***adjusted p-values < 0.001.

Next, we investigated whether the top SNP loci identified in our study influenced the expression of nearby genes, specifically as cis-expression quantitative trait loci (eQTLs) (**Figure 5B, Supplementary Table 8B**). rs3747973 was found to influence the expression of *RAB29*, which did not show significant differences between populations. Similarly, rs34311866 was associated with the expressions of *GAK*, *DGKQ*, and *IDUA. DGKQ* showed significant differences between AFR and the other populations, while *IDUK* exhibited varying expression differences across multiple population pairs. Two SNPs, rs2517680 and rs1846190, were linked to the regulation of other HLA-class genes, including *HLA-DRB6*, *HLA-DRB1*, *HLA-DQA2*, and *TAP2*. The widespread expression variability observed in these loci underscores the importance of HLA-related genes in PD pathogenesis. In addition, rs56328224 influenced the expression of approximately ten genes, including *KANSL1-AS1* and *KANSL1*, which displayed expression variability across multiple population pairs. Fine-mapping analysis using SuSiE further revealed that this SNP regulates *NSFP1* and *ARL17B*, both of which exhibited significant differences in expression between SAS and EAS, AFR, and AMR populations (**Figure 5C, Supplementary Table 8C**). Finally, rs2092563 was associated with regulating 11 genes including *TEF*, *PMM1,* and *MEI1*, with expression variability observed across populations. (**Figure 5B, Supplementary Table 8B**).

Based on the fastQTL analysis, the rs9393839 SNP identified in the meta-analysis affected the expression of five genes. Among them, *ZNF391*, *ZNF165*, and *ZSCAN31* all zinc finger proteins, showed differential expression across populations. *ZNF391* expression differed significantly between EAS and multiple populations. *ZNF165* expression varied significantly between AMR and other populations, as well as between EUR and EAS. *ZSCAN31* expression was significantly different between EAS and EUR/AFR, as well as between AMR and AFR (**Supplementary Table 8E**).

Complimentary to the MAGE dataset, gene and tissue enrichment analysis with MAGMA and FUMA identified associations with 75 genes (**Supplementary Table 9**). Fifteen of these genes overlapped with results from the MAGE dataset. Tissue expression analysis revealed enrichment in skeletal muscle, whole blood, substantia nigra, and left ventricle of the heart, though these findings did not achieve statistical significance, likely due to the limited sample size in our study (**Supplementary Figure 16**). Integrating the MAGE dataset from 1000G populations with our key loci revealed substantial expression differences across global populations, providing insights into population-specific gene expression and its potential impact on PD pathogenesis.

## Discussion

We present the findings of the largest genetic survey of PD conducted to date in the Indian population. Our study revealed several key observations: 1) Heritability estimates indicate a substantial genetic burden of PD in the Indian population with an 8% higher genetic load compared to the European population (30% vs 22%)^28^. 2) Common genetic loci implicated in European populations, including *SNCA*, *TMEM175*, *LRRK2,* and *MAPT* are also associated with PD in the Indian population. Most loci identified in the European meta-analysis exhibit consistent directionality, suggesting a similar disease risk continuum across these populations. 3) The risk conferred by *LRRK2* in the Indian population is independent of G2019S, and Asian-specific *LRRK2* risk variants (G2385R, and R1628P) are rare. 4) Unlike the common *GBA1*-driven mutations in European populations, the p.E326K and p.T369M variants emerge as significant risk factors for PD in the Indian population. 5) The H2 haplotype appears protective, whereas most H1-clade haplotypes show no association with PD in the Indian population. 6) The loci identified exhibit significant expression variability across global populations, underscoring distinct mechanistic underpinnings for PD in different ethnic groups.

Despite allelic heterogeneity, our findings reveal substantial overlap in common genetic variability at key risk loci previously implicated in PD, underscoring the importance of loci such as *NUCKS1, ITPKB, TMEM175,* and *SNCA* in disease pathogenesis. However, our global expression analysis suggests that the underlying mechanisms may vary across populations The *SNCA* locus exhibits a consistent association across diverse populations, as previously reported^30^. Notably, three independent signals are known to drive this association at the *SNCA* locus^31,32^. In our study, two of these variants reached genome-wide significance, while the third (rs287004) showed suggestive evidence with a p-value of 2.64 × 10⁻⁴, reinforcing the role of *SNCA* in the Indian population.

Our findings expand the mechanistic landscape of PD by implicating newly identified loci in lysosomal-autophagy pathways, vesicular trafficking, and lipid membrane dysregulation. For instance, *ATP10A*, a member of the P4-ATPase family, plays a critical role in lipid translocation across bilayer membranes, influencing processes such as signal transduction, cell division, and vesicular transport^33,34^. A previous study showed that genetic variability within the *ATP10A* gene is associated with insulin resistance in nondiabetic African American patients. Insulin resistance is known to contribute to a severe phenotype and disease progression in PD^35^. The role of lipid aggregation in PD remains unclear, particularly in the context of *GBA*-linked mutations. Glucosylceramide (GlcCer) accumulation in the brain and plasma of PD patients has been documented^36,37^, and conserved domains within the transmembrane 4 region of P4-ATPases, such as *ATP10A* and *ATP10D*, serve as critical substrates for GlcCer translocation^38^. These findings suggest that genetic variability at the *ATP10A* locus could provide new strategies to elucidate the role of GlcCer in PD.

*MED13* has been identified as an *SNCA* modifier, with studies showing that its upregulation can induce *SNCA*-linked neurodegeneration in the *Drosophila* model^39^. Located 16 Mb downstream of the *MAPT* locus^40^, *MED13* has also been linked to compensatory glycolysis mechanisms that counteract neurodegeneration^39^. Similarly, *MICD* is a pseudogene, and emerging evidence suggests that the processing of pseudogenes into short interfering RNAs can regulate coding genes through the RNA interference (RNAi) pathway in neurodegeneration^41^. Furthermore, *EP300* has been implicated in aberrant tau secretion in neuronal models, highlighting its role in tauopathy^42^. Variability in these loci may offer critical insights into the dysregulation of lysosomal, vesicular, and lipid-associated pathways, advancing our understanding of PD pathogenesis. A GWAS on Alzheimer’s disease identified the *EP300* gene as one of the top loci influencing stage-specific effects^43^. The loci identified in our meta-analysis underscore varied functions ranging from the role of pseudogenes (*RLP8L1*) to regulating the adaptive immune response (*TEC*) in PD. Finally, the Down Syndrome Cell Adhesion Molecule (*DSCAM*) gene, which belongs to a superfamily of cell adhesion molecules, encompasses the DS critical region and is involved in the development of the central and peripheral nervous system.

The loci identified in our study expand the genetic map of PD by highlighting the diverse roles of these genes and offering new mechanistic insights into how dysfunctions in membrane formation, immune dysfunction, membrane transport and dysregulation of tau secretion along with the propagation via EP300 might influence neurodegeneration in PD^42^. Gene expression analysis highlights the *RAB29* as an eQTL locus for the *NUCKS1* locus, an observation consistent with previous studies^19,44^. RAB proteins, as members of a superfamily, act as molecular switches cycling between GDP-bound inactive and GTP-bound active forms^45^. Notably, *RAB29* has been shown to act upstream of *LRRK2*, regulating its kinase activity^45^. It would be interesting to follow up on how genetic variability within these genes modulates *LRRK2* activity in the Indian population independent of the most common kinase and/or ROC-COR domain mutations.

While this study provides a comprehensive understanding of the genetic architecture of PD in the Indian population, significant challenges persist, particularly in data-sharing infrastructure. These limitations stand in stark contrast to the well-established research ecosystems in Europe and the USA. New strategies are necessary to better integrate research efforts from countries like India into the global scientific landscape. Supporting the development of local infrastructure, and human resource growth, and encouraging the harmonization of data processes globally would contribute to a more comprehensive global understanding of Parkinson’s disease. Indeed, efforts are being made to address this disparity in PD research^14,46^. Over the past decade, the Indian government, through the Indian Council of Medical Research (ICMR) and other funding agencies, has made strides in promoting collaborative work and developing resources and infrastructure in this direction (https://epms.icmr.org.in/). In parallel, investment from the private sector along with active support from international funding agencies such as MJFF and GP2, is driving momentum for PD research in India. Through this study, we have provided a mechanism which will allow researchers to collaborate and advance PD research and allow the Lux-GIANT consortium to actively engage in global initiatives (**Supplementary Figure 17**).

Our study has limitations, primarily the absence of an independent replication cohort. Efforts are already underway to recruit PD patients across India as part of a second phase. To address the potential for false-positive findings, we employed a single-stage study design that ensured a well-powered GWAS. Moreover, the consistent directionality of the effect estimates for key loci, as observed in the European population and the meta-analysis, reduces the likelihood of spurious associations.

Importantly, this study established a well-characterized PD biobank, providing a critical resource for advancing patient-centric approaches to Parkinson’s disease research and treatment in India. Our findings highlight the significant genetic burden of PD in the Indian population and underscore the need for continued engagement with healthcare providers and patient organizations to improve awareness and care. In addition, the platform developed as part of this work enables Indian researchers to access valuable data, thereby promoting the expansion of PD research and facilitating deeper exploration of population-specific variability on PD pathogenesis.

## Acknowledgements

The authors wish to thank the patients, healthy volunteers and members of the Lux-GIANT consortium who have contributed to this effort. We acknowledge the Indian Council of Medical Research (ICMR) in facilitating the Lux-GIANT consortium to conduct the current study. This project was supported by the Global Parkinson’s Genetics Program (GP2; https://gp2.org). GP2 is funded by the Aligning Science Across Parkinson’s (ASAP) initiative and implemented by The Michael J. Fox Foundation for Parkinson’s Research (MJFF). For a complete list of GP2 members see doi.org/10.5281/zenodo.7904831. This project was also supported by the Michael J. Fox Foundation for Parkinson’s Research Grant ID MJFF-017473.

## Author contributions

These authors contributed equally: Asha Kishore, Ashwin Ashok Kumar Sreelatha, and Amabel M. M. Tenghe.

Study design and project coordination: Manu Sharma, Asha Kishore,

Analysis team: Amabel M.M. Tenghe, Ashwin Ashok Kumar Sreelatha, Luis-Giraldo Gonzalez-Ricardo, Kandadai, Saeideh Khodaee, Dan Vittale, Mike Nalls

Sample collection and management: Manu Sharma, Asha Kishore,Rupam Borgohain, Divya Kalikavil Puthanveedu, Roopa Rajan, Madhusoodanan Urulangodi, Pramod Kumar Pal, Rukmini Mridula Kandadai, Ravi Yadav, Sahil Mehta, Hrishikesh Kumar, Niraj Kumar, Prashanth Lingappa Kukkle, Soaham Dilip Desai, Kuldeep Shetty, Pettarusp Wadia, Annu Aggarwal, Pankaj Agarwal, Mirza Masoom Abbas, Gurusideshwar Mahadevappa, Wali, Syam Krishnan, Divya Madathiparambil Radhakrishnan, Nitish Kamble, Achal Kumar Srivastava, Vivek Lal, Teresa Maria Costa Ferreira, Manas Chacko, Cibin Thoppil Raghavan, Gangadhara Sarma, Justin Solle, Brian Fiske, Amiya Thalakkatttu, Divyani Garg, Jens Krüger, Peter Lichtner, Dan Vittale, Mike Nalls, Cornelis Blauwendraat, Andy Singleton, Monojit Debnath, Swagata Sarkar, Sabbir Ansari, Sachin Adukia, Pravi Vidyadharan, Kanthimathi R, Santhi CKV, Tazeem Fathima Syed, Sindhuja Mohareer, Global Parkinson’s Genetics program (GP2), Lux-GIANT consortium. All authors contributed to the final version of the manuscript.

## Supplementary information

a) Supplementary_Figures_1-17.docx

b) Supplementary_Tables_1-10.xlsx

## Methods

### Participants

A detailed description of the recruitment process is provided elsewhere^1^. In brief, 18 centres across India participated in this study. All cases were diagnosed and enrolled by movement disorder specialists to ensure diagnostic accuracy for Parkinson’s disease (PD). Before inclusion, participants underwent a thorough medical history review and standard neurological examination. PD diagnoses were made based on the United Kingdom Parkinson’s Disease Society Brain Bank diagnostic criteria^2^. Subjects with severe cognitive dysfunction or active psychosis, which impaired their ability to provide written informed consent, as well as those exhibiting red flags suggestive of atypical parkinsonism, were excluded from the study. Healthy controls, who were gender-matched and from the same geographic and ethnic background as the cases, were recruited through advertisements posted on hospital campuses. Individuals with a family history of PD, tremor, or other neurodegenerative diseases were excluded from the control group. Finally, the Research Electronic Data Capture (REDCap) platform was implemented for our Lux-GIANT cohort to harmonize the PD data^3^. REDCap is a secure platform in which all survey data are recorded and stored on a database server at SCTIMST, following stringent security protocols. Additionally, sensitive data are pseudonymized to ensure extra protection.

### Genotyping

Genotyping was conducted at MedGenome Inc., Bangalore, India. Genomic DNA was extracted from blood samples using a genomic DNA extraction kit. DNA quantity was assessed, and quality was verified via agarose gel electrophoresis. The genotyping was performed using the Global Diversity Array + NeuroBooster chip (Illumina Inc.), which includes probes for approximately 1.89 million markers. DNA samples meeting the required quality and quantity standards were amplified, enzymatically fragmented, and precipitated with isopropanol. Afterwards, the DNA was re-suspended and loaded onto bead chips within hybridization chamber inserts for further analysis. The bead chips were placed into hybridization chambers and incubated in an Illumina Hybridization oven at 48°C for 16-24 hours. Following hybridization, the chips were washed and prepared for single-base extension. After extension, the chips were stained with fluorescent dyes: biotin for G and C nucleotides, and DNP for A and T nucleotides. The chips were then scanned using the iScan system. Genotype data for 12,949 samples were generated using the NeuroBooster Array (NBA), which includes approximately 1.89 million markers. The raw data (IDAT files) were processed using the Illumina Array Analysis Platform Genotyping Command Line Interface to generate genotype calls (GTC). These GTC files were subsequently converted to Variant Call Format (VCF) using the GTC to VCF tool from Illumina. The NBA array manifest file and the GRCh38 FASTA files were provided as inputs for this conversion process. Sample-specific VCF files were produced through this pipeline, compressed with the bgzip tool, and their corresponding MD5 sums were generated.

### Quality Control

The raw genotype files were converted into PLINK binary formats, and quality control was conducted using PLINK^4^. Genotyping quality was assessed by excluding samples with a call rate below 98%, leading to the removal of 9 samples. Gender discrepancies were evaluated using heterozygosity on chromosome X, with the gender assigned based on the inbreeding coefficient (*F*) calculated from chromosome X genotypes: males (*F* > 0.8) and females (*F* < 0.2). Samples with ambiguous *F* values or discrepancies between reported and genotyped sex were flagged and excluded, resulting in the removal of 40 samples. Heterozygosity on autosomes was analyzed to identify potential genotyping errors or sample contamination, and 153 samples exceeding ±3 standard deviations from the mean heterozygosity were excluded.

To address non-reported familial relationships, an assessment of relatedness was performed by estimating a kinship matrix using the KING-robust estimator algorithm. Closely related individuals, identified by a kinship coefficient threshold of > 0.354, were resolved by randomly excluding one individual from each pair. This step resulted in the removal of 362 samples and ensured the independence of the remaining individuals for downstream analyses.

Population structure was examined by merging the study dataset with genotyped variants from the multi-ethnic 1000 genomes phase 3 reference panel^5^, yielding 618,666 variants. The merged dataset was filtered for variants with a minimum minor allele frequency (MAF) of 1% and pruned for linkage disequilibrium (LD) using a 50-variant window, a 5-variant step, and R^2^ < 0.2 as parameters. Principal component analysis (PCA) was performed, and the first two principal components were projected to identify ethnically mismatched samples. Outliers were defined as samples exceeding +4 standard deviations from both the mean of South Asian reference samples and the mean of the study cohort. A total of 405 outliers were excluded. Twelve controls identified to have a PD family history were also excluded. To further investigate population structure, a Uniform Manifold Approximation and Projection (UMAP) was computed using the first 10 principal components (PCs) computed with PLINK as described above. The UMAP projections were visualized in Python to ensure consistency with PCA results.

For SNP (single nucleotide polymorphism) quality control, only SNPs genotyped in at least 99% of the final dataset were retained, resulting in the removal of 5,100 SNPs. Variants with a minor allele frequency (MAF) below 5 × 10⁻⁸ were excluded (448,403 SNPs removed). Additional filtering was performed separately for males and females: SNPs with a marker call rate below 98% for females (102,617 SNPs removed), males (9 SNPs removed), or cases and controls with a call rate below 1 × 10⁻^5^ (12,617 SNPs removed) were excluded. SNPs showing significant deviation from Hardy-Weinberg equilibrium in controls (*P* < 5 × 10⁻⁸, 8,643 SNPs removed) were also excluded. After applying all quality control filters, the final dataset consisted of 1,290,001 high-quality SNPs across 11,170 individuals, which were used for downstream analyses.

### Imputation

After quality control, we used the imputation preparation and checking tool available at https://www.well.ox.ac.uk/~wrayner/tools/HRC-1000G-check-bim-v4.3.0.zip to identify and correct inconsistencies, including allele mismatches, strand orientation errors, allele frequency discrepancies, and problematic palindromic SNPs. Imputation was performed using the Michigan Imputation Server, with the 1000 Genomes reference panel and the GRCh38/hg38 human genome assembly. All SNPs with accuracy (R^2^) of imputation ≥ 0.3 were retained for subsequent analysis.

### Genome-wide association testing

Imputed SNPs were included in the association analysis if they fulfilled the following filtering criteria: call rate > 0.1, MAF > 0.01, and no significant deviation from HWE (*P* < 5 × 10⁻^8^. The final genotype dataset consisted of 8,414,146 SNPs across 11,170 individuals (4,806 cases and 6,364 controls), with only autosomal variants retained. Before the GWAS, the imputed dataset was LD pruned as previously described and used to compute PCs and a genomic relation matrix (GRM). Association analyses were conducted using the case-control logistic regression model in PLINK, with sex and the first 10 PCs included as covariates. A Bonferroni-corrected significance threshold of *P* < 5 × 10⁻⁸ was applied, with variants showing *P* < 1 × 10⁻⁶ classified as suggestive. To correct for residual population stratification or cryptic relatedness, genomic control (GC) was applied by adjusting the test statistics using the genomic inflation factor (λGC), calculated as the ratio of the median observed chi-square statistic to its expected value under the null hypothesis. Post-adjustment, quantile-quantile (QQ) plots and recalculated λGC confirmed effective correction. Independent signals were identified through approximate conditional analyses using the COJO method in GCTA^6^, conditioning on GWAS-significant SNPs as covariates during restricted maximum likelihood (REML) analysis. Signals were considered independent if the conditioned p-value was < 5 × 10⁻⁸.

We also tested SNP effects with a mixed model using the fastGWA-GLMM method in GCTA, which incorporates the GRM as a random effect to account for cryptic relatedness, with sex and the first 10 PCs as covariates, and the results were further refined using COJO. To harmonize our summary statistics with ongoing GWAS studies in the GP2 consortium, additional analyses were performed using GenoTools^7^, yielding consistent results. Furthermore, we developed an Integrated Downstream Analytical platform for genomic analyses (IDEAL-GENOM) for the Lux-GIANT cloud, that combines standard quality control, GWAS and downstream analysis in one suite.

### SNP-based heritability

To estimate a larger portion of the additive genetic variance not typically explained by GWAS, we assessed SNP-based heritability by stratifying the SNPs into three datasets using PLINK: 1) all imputed SNPs used in the GWAS, 2) SNPs located within ± 1MB of independent associated SNPs identified in the GWAS for the PD case-control dataset, 3) SNPs not located within ±1MB of the independent associated SNPs. The 13 lead SNPs in Table 1 were used to define the PD regions for the second and third datasets. For each of the datasets, the genotypes were LD pruned as previously described, and a GRM was computed. The GRMs were then used in REML analysis to estimate heritability in GCTA. To account for ascertainment, variance estimates were transformed from the observed scale to the underlying scale. We estimated heritability for PD case-controls, age at onset, early age at onset and late age at onset for each of the stratified data. Cases with an age at onset below 50 years were classified as early-onset, while those with an age at onset above 50 years were categorized as late-onset. Considering the age-related increase in PD prevalence, we applied standardized disease prevalence rates for PD, adjusted for age and gender, as reported in previous studies^8–11^.

### Meta-Analysis

We performed a meta-analysis of GWAS summary statistics from our study and publicly available multi-ancestry GWAS data using the genome-wide association meta-analysis (GWAMA) software^12^. The primary analysis employed the fixed-effect inverse-variance weighted model, which assumes a consistent true effect size across studies. Summary statistics were obtained for a published multi-ethnic cohort consisting of 611,485 individuals of European, East Asian, and Hispanic ancestry^13,14^. After harmonizing allele orientations and excluding ambiguous SNPs across the multi-ethnic cohort and our study population, 7,097,037 variants were included in the meta-analysis.

The resulting association statistics were annotated and filtered to identify genome-wide significant loci (*P* < 5 ×10^−8^) and loci suggestive of association (*P* < 1 × 10^−6^). Conditional analyses were performed using GCTA to pinpoint independent loci, incorporating pairwise LD from the multi-ethnic 1000 Genomes Phase 3 reference panel. Independent SNPs were defined as those retaining genome-wide significance (*P* < 5×10^−8^). In addition, we performed a meta-analysis using the random-effects model, which accounts for potential heterogeneity by assuming random variation in effect sizes across studies.

### MAPT haplotype analysis

To investigate the role of MAPT haplotypes in PD following the GWAS, we defined haplotypes using six variants: rs1467967, rs242557, rs3785883, rs2471738, rs8070723, and rs7521^15^. The tagging variant rs8070723 was used to distinguish between the major H1 and minor H2 haplotypes of the MAPT locus, where the major allele corresponds to the H1 haplotype, and the minor allele corresponds to the H2 haplotype.

Imputed genotype data for these six variants were extracted to define the MAPT haplotypes. Associations between haplotypes and PD risk were evaluated using the haplo.stats package in R^16,17^. Haplotype probabilities were estimated using the haplo.em function, which employs an expectation-maximization algorithm to infer haplotypes from genotype data. For each individual, the expected number of copies of a given haplotype was calculated based on the estimated probabilities. Logistic regression models were used to assess the association between haplotype copy number and PD risk, adjusting for sex as a covariate. Haplotypes occurring in < 1% of individuals were excluded from the analysis.

Statistical significance was set at a Bonferroni-corrected threshold to account for multiple comparisons. Associations with p-values < 0.0031 (16 tests, corresponding to 16 haplotypes with ≥ 1% frequency) were considered statistically significant. Odds ratios with corresponding 95% confidence intervals were calculated for each haplotype, representing the effect of an additional copy of the given haplotype on PD risk.

### FUMA and MAGMA

Gene and tissue enrichment analyses were performed using MAGMA v1.08 on the FUMA platform (v1.6.0). P-values from the GWAS were used to evaluate associations at both the gene and gene-set levels. In the gene-based analysis, 10,678 gene sets from the MSigDB database (version 7.0) were tested, with multiple comparisons corrected using the False Discovery Rate (FDR). For tissue-specific enrichment, the GWAS dataset was analyzed to assess the enrichment of associations among genes expressed across 53 tissue types from the GTEx v8.0 database. This integrated approach enabled a thorough investigation of how gene expression patterns and tissue-specific expression contribute to the associations identified in the dataset.

### Polygenic risk prediction

We used summary statistics of published GWAS or meta-analyses for PD in European (EUR) and Indian (IND) populations as base data^13,18^. The EUR base summary statistics consisted of 17,510,617 variants from 578,413 individuals, while the IND base comprised 588,081 variants from 2,050 individuals. The software PRSice-2^19^ was used to calculate polygenic risk scores (PRS) using each of the base datasets. The PRS were calculated by computing the sum of risk alleles corresponding to the base phenotype (Parkinson’s disease) in each individual, weighted by the effect size estimate derived from the base GWAS. We clumped SNPs at different thresholds and a 500kb window with r^2^ of 0.1 was the best fitting and performance model for the EUR base set, while the IND base set performed best at 500kb window with r^2^ of 0.5 and 250kb window with r^2^ of 0.5. We also tested different *P-*value thresholds (*P_T_*), ranging from 1 to 5 x 10^-8^ for selecting clumped SNPs to be included in the final PRS model. The *P*-value threshold which accounted for the highest proportion of variance (*R*^2^) was selected as the best PRS for PD. The explained variance (Nagelkerke’s *R*^2^) was derived from a generalized linear model in which PD status (target phenotype) was regressed on each PRS, adjusting for covariates (sex, and first ten PCs computed from the genotype data in PLINK). An incremental *R*^2^ was computed from each model by PRSice-2 and plotted against the *P_T_*. The *R*^2^ is the difference between the *R*^2^ of the full model (*PD status ∼ PRS + Covariates*), and *R*^2^ of the null model (*PD status ∼ Covariates*). We used a conservative prevalence of 0.5% as in previous studies for the PRS computation^20,21^.

To test to what extent PRS can be used for risk stratification of individuals with PD, we categorized the individuals into quartiles according to the PRS and compared the odds ratio (OR) across quartiles. The bottom quartile was used as a reference and compared to the other quartiles, using logistic regression. To test the prediction performance of the models, receiver-operating curves (ROC) were derived and the corresponding area under the curves (AUC) was computed. DeLong’s test in the R packag*e pROC*^22^ was applied to compare the AUC of the full models and covariate models, allowing assessment of the additional specific contribution in prediction performance attributable to the PRS.

### PD loci and gene expression

Quantified gene expression data from the MAGE dataset, generated from lymphoblastoid cell lines derived from 731 individuals in the 1000 Genomes Project, were used for analyses^23^. This dataset was used to identify genetic variants associated with the expression levels of nearby genes (cis-eQTLs). First, FastQTL analysis^24^ was performed to identify SNPs significantly associated with gene expression levels. This method employs a regression-based approach to detect genetic variants influencing expression, even in large datasets, by leveraging expression quantitative trait loci (eQTL) mapping. Following this, fine mapping was carried out using “Sum of Single Effects” (SuSiE)^25,26^ to pinpoint credible sets of variants responsible for driving the QTL signals. The SuSiE model applies a Bayesian framework to identify multiple causal variants in genetic associations, modelling each variant’s effect as a sparse vector with a single non-zero component. It calculates posterior inclusion probabilities (PIPs) for each variant, quantifying the likelihood that a variant is causal, and constructs high-confidence credible sets containing at least one causal variant. Using SuSiE for fine-mapping in eQTL studies allows for the discovery of multiple independent causal signals per gene, providing small, high-resolution credible sets with minimal correlation among variants.

For the top SNP loci identified in our study, we utilized data from a previous analysis that reported 42 genes whose expression is significantly associated with these SNPs using the FastQTL method^23^. Additionally, three genes were found to be affected by a causal variant (SNP: rs56328224) through the SuSiE approach^23^. We extracted the expression levels of these genes, along with those identified as nearby to the SNPs from the processed data and compared their expression across different superpopulations.

To assess the statistical significance of differences in median expression levels between two superpopulations, a permutation test was used to calculate p-values. First, the difference in medians between the two groups was calculated. To generate the null distribution, data from both groups were pooled, and the labels corresponding to the groups were randomly shuffled, disrupting any inherent association between individuals and their respective groups. This process was repeated 100,000 times to build the null distribution for each pairwise comparison. The observed test statistics were then compared against the null distribution, with the p-value for each comparison determined by calculating the proportion of permuted differences greater than or equal to the observed median difference. To account for multiple comparisons, the p-values were adjusted using the Benjamini-Hochberg (FDR) method. A p-value of less than 0.05, after adjustment, was considered statistically significant for each pairwise comparison.

### Data Management and Sharing

All participating Indian nodal centres and sub-centres in this study are collecting consented and pseudonymized data locally within secure and access-controlled environments. Identifiable information for each study participant is stored separately at each site, while pseudonymized data is transferred to a central data hub. Access to the clinical data is restricted to authorized investigators from the respective sites, who are responsible for handling the clinical information (clinical core). This setup complies with both the European General Data Protection Regulation (GDPR) and the Indian Personal Data Protection Bill (PDP) 2019. The pseudonymized clinical data, along with the corresponding genetic data, is accessible to all Lux-GAINT partners through the “data core” platform hosted at the de.NBI Cloud Tübingen (https://cloud.denbi.de). The data core provides a secure environment for storing genetic data and offers processing capabilities for its analysis. Depending on the requirements, volume size, and anticipated processing steps, different storage models are available. Storage volumes based on the Quobyte (https://www.quobyte.com) storage system are secured with individual user certificates, enabling fine-grained access control. A similar setup, providing secured volumes as directly attached volumes, is available using the Ceph (https://ceph.io) storage system. Despite being technologically distinct, both solutions share the feature that volumes reside within separate VLAN/VXLAN networks, preventing external access. As a result, virtual machines used for processing LUX-Giant genomic data must be specifically configured to allow access. Additionally, certain volumes can be exposed as S3 buckets, maintaining tight access control, which is commonly used for reference data or data sharing.

The organizational and technical process, beginning with the project application, is illustrated in Supplementary Figure 18. For additional details, please refer^1^. The established system infrastructure ensures a secure environment for handling and processing sensitive medical data in a controlled and responsible manner, utilizing cloud resources. The de.NBI Cloud Tübingen undergoes regular security audits, including penetration testing, and its information security management system is certified according to ISO 27001 standards.

## Data availability

Summary statistics from the GWAS and meta-analysis are available via Zenodo (https://doi.org/10.5281/zenodo.14726796) and GP2 tier 1 at the AMP-PD portal (https://amp-pd.org/), and are publicly available as of the date of publication.

## Code availability

All code generated for this article, and the identifiers for all software programs and packages used, are available on GitHub https://github.com/GP2code/India-metaGWAS and were given a persistent identifier via Zenodo (10.5281/zenodo.14755663)

