## Supplementary Figure for "Deciphering the Genetic Architecture of Parkinson’s Disease in India"

**Supplementary Figure 1:** Q-Q plots from GWAS

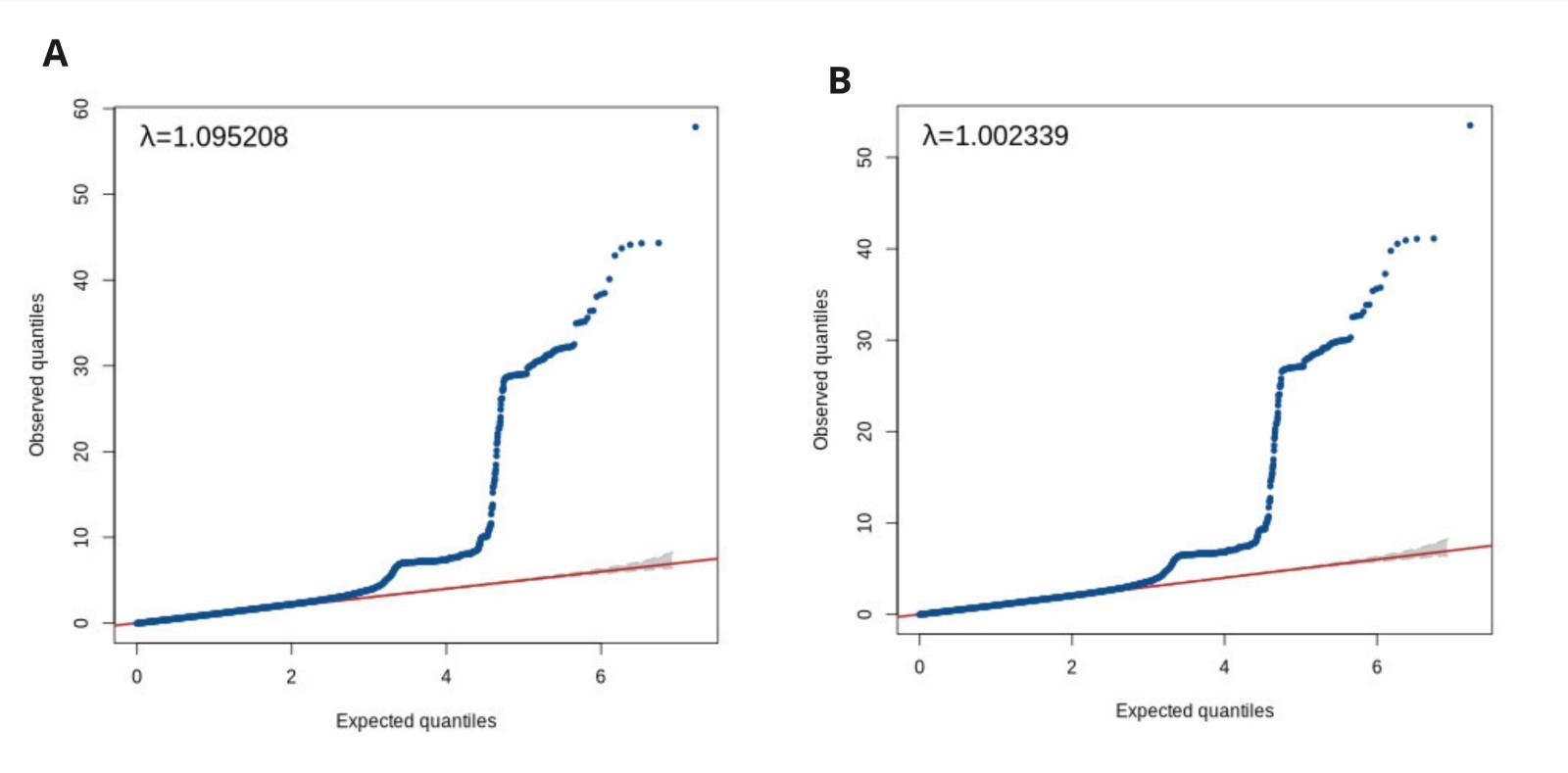
(A) unadjusted and adjusted (B) for genomic control for PD GWAS with a generalized linear model using PLINK.

**Supplementary Figure 2-13:** Locus Zoom plots of genome-wide significant loci in the Indian population

2. LocusZoom plot of the association signal of the NUCKS1 (rs3747973)

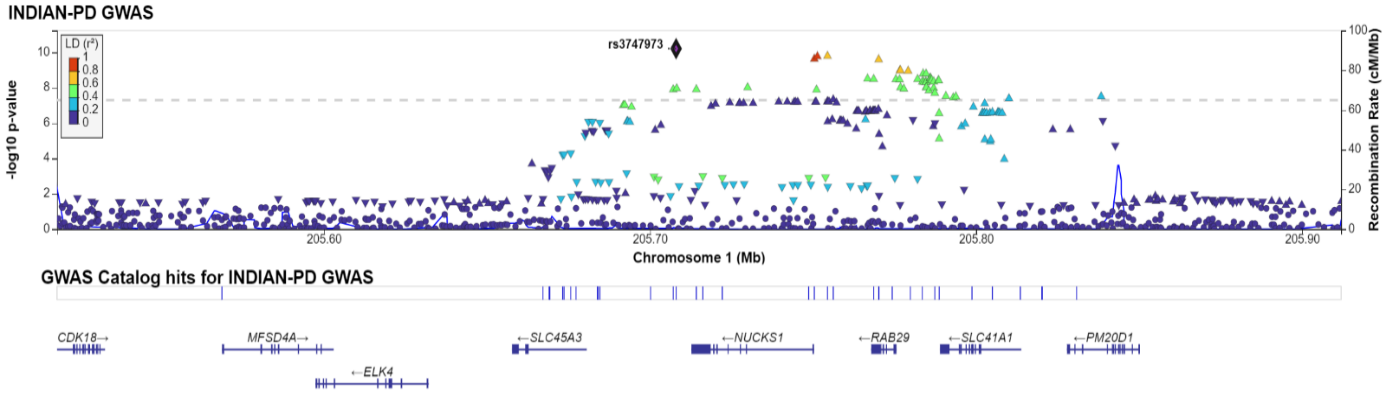

3. LocusZoom plot of the association signal of the ITPKB (rs74990530)

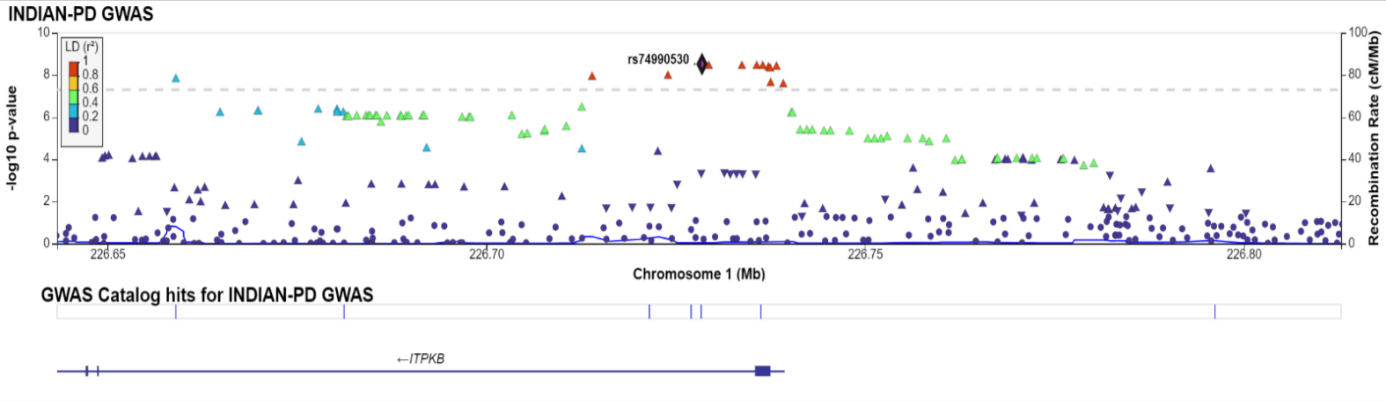

4. LocusZoom plot of the association signal of the TMEM175 (rs34311866)

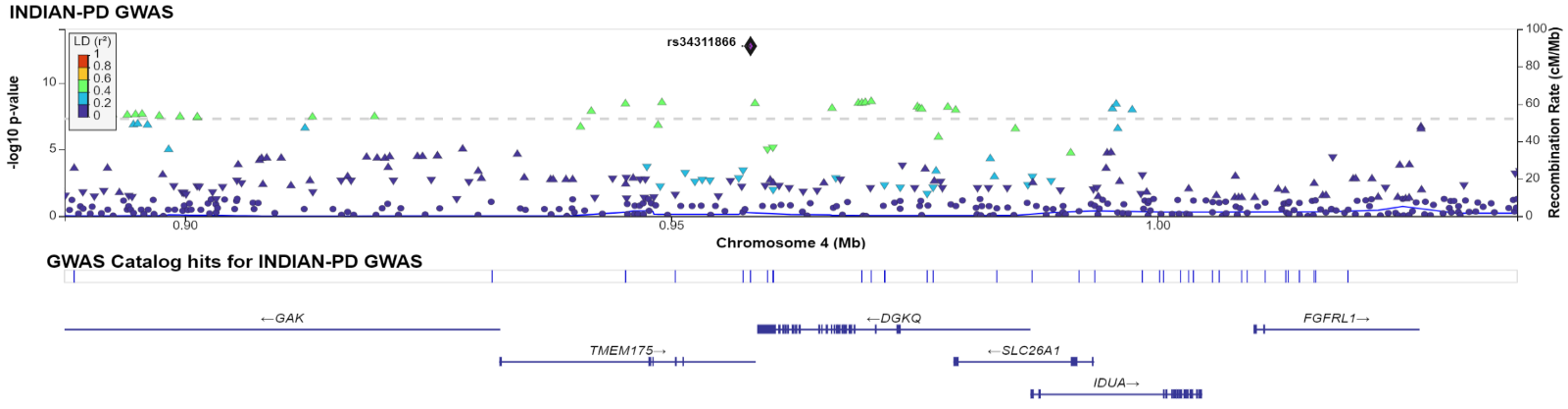

5. LocusZoom plot of the association signal of the SNCA (rs356182) and (rs7681440)

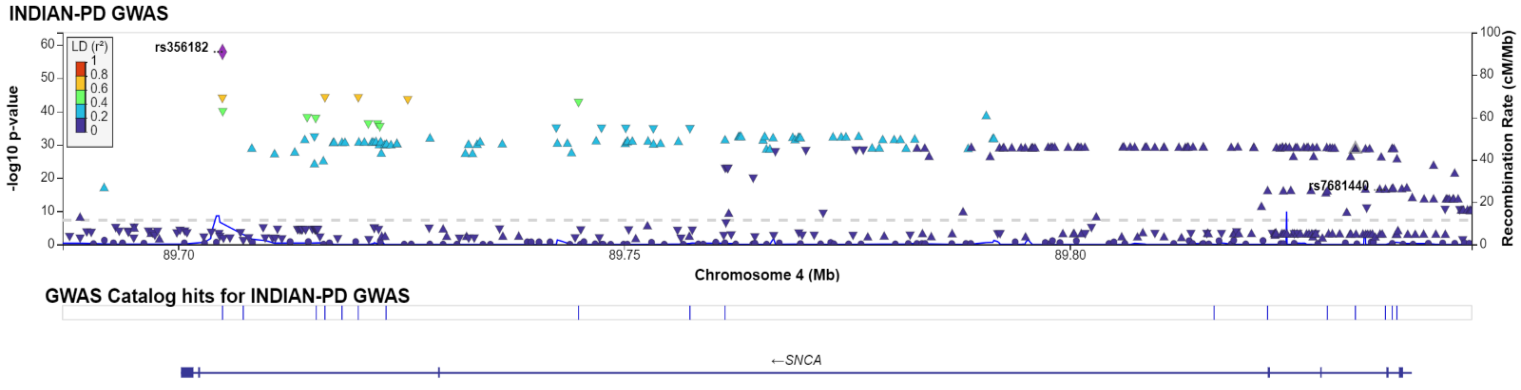

6. LocusZoom plot of the association signal of the HLA-DQA1 (rs1846190)

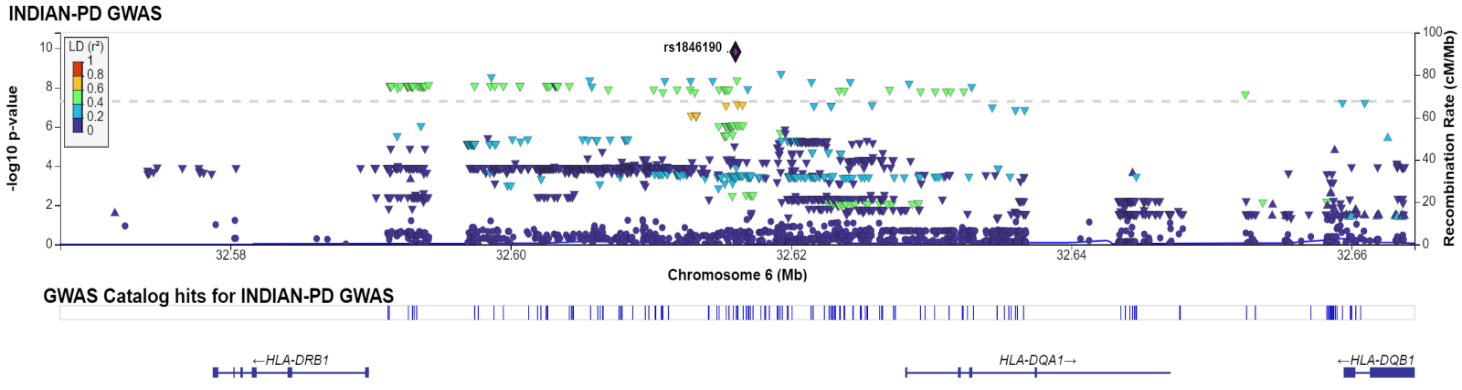

7. Locus zoom plot of the association signal of the MICD (rs2517680)

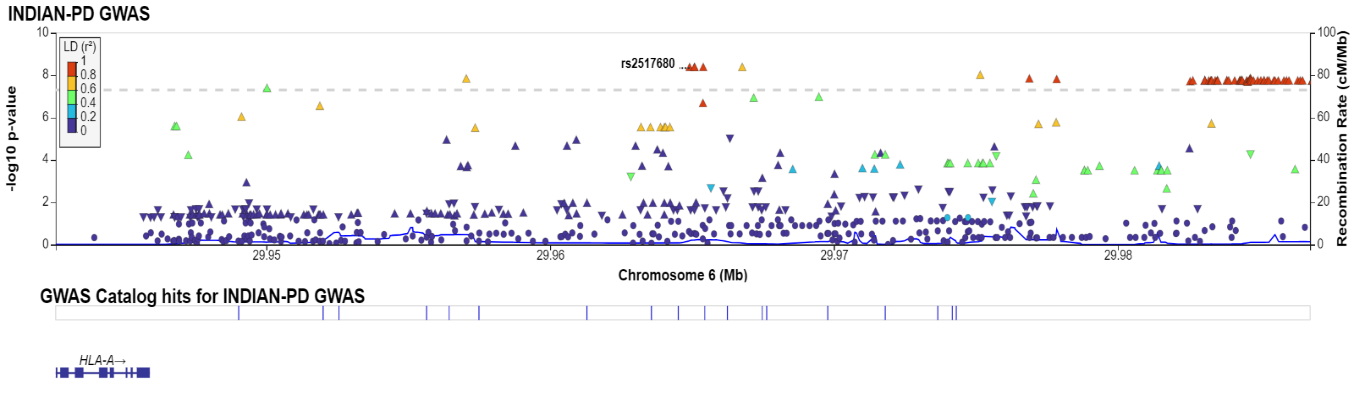

8. Locus zoom plot of the association signal of the RNF141 (rs490994)

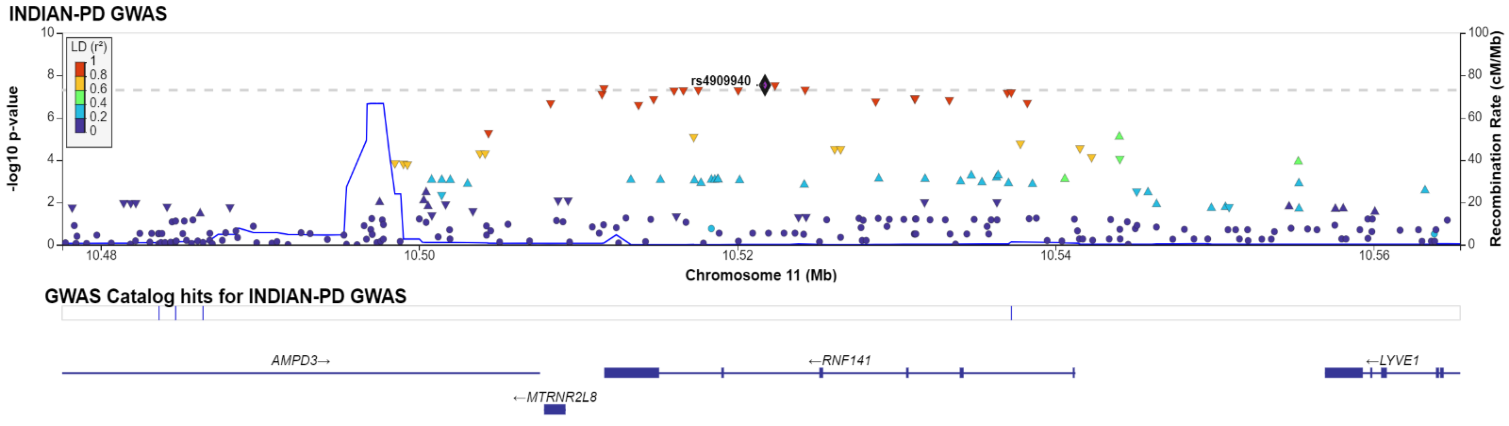

9. Locus zoom plot of the association signal of the ATP10A (rs528813377)

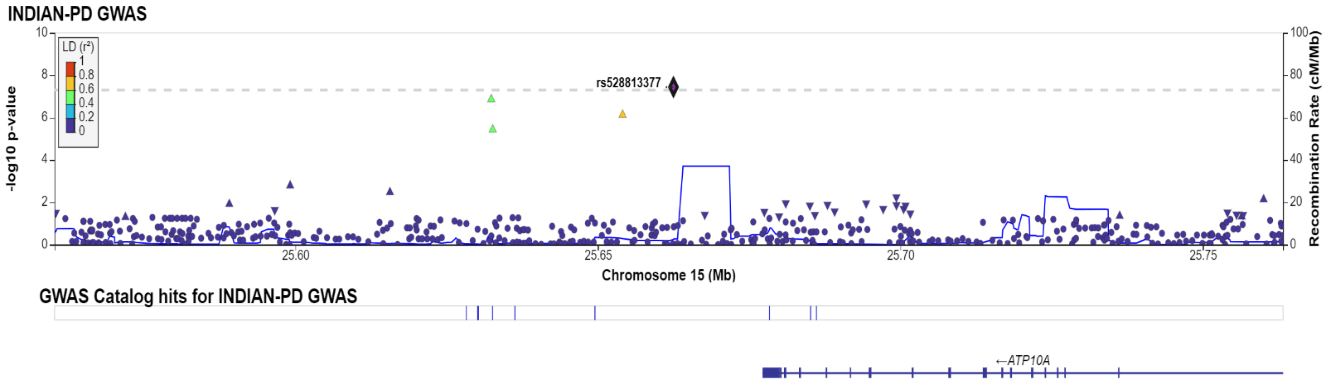

10. Locus zoom plot of the association signal of the PLEKHM1 (rs56328224)

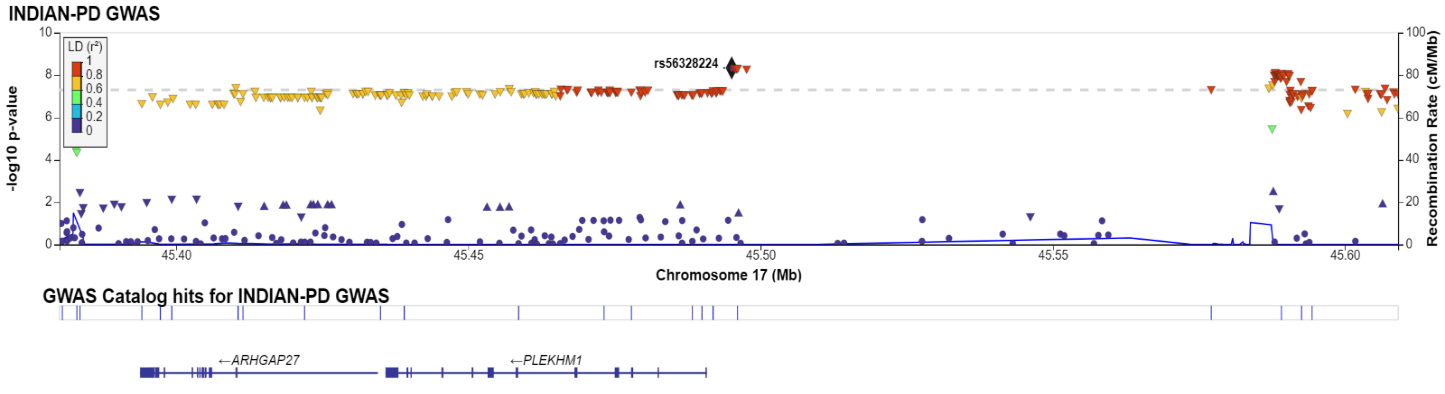

11. Locus zoom plot of the association signal of the MED13 (rs72843781)

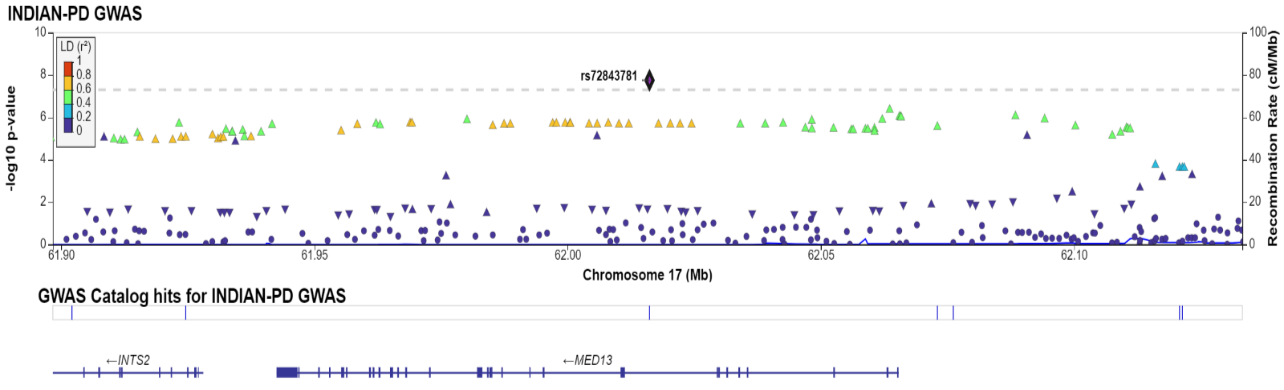

12. Locus zoom plot of the association signal of the RIT2 (rs8087199)

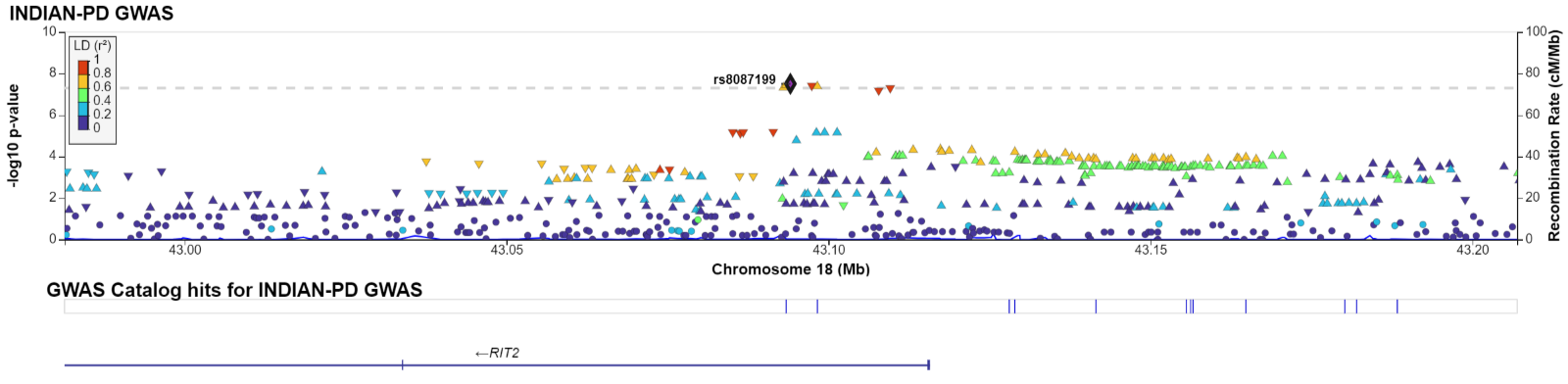

13. Locus zoom plot of the association signal of the EP300-AS1 (rs2092563)

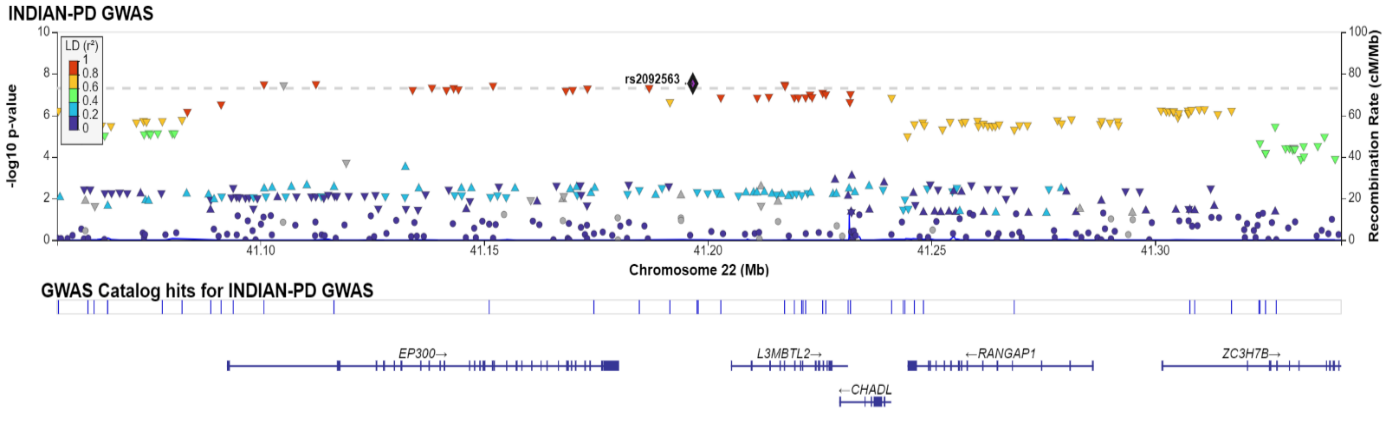

**Supplementary Figure 14.** Effect estimates Indian GWAS vs European meta-analysis

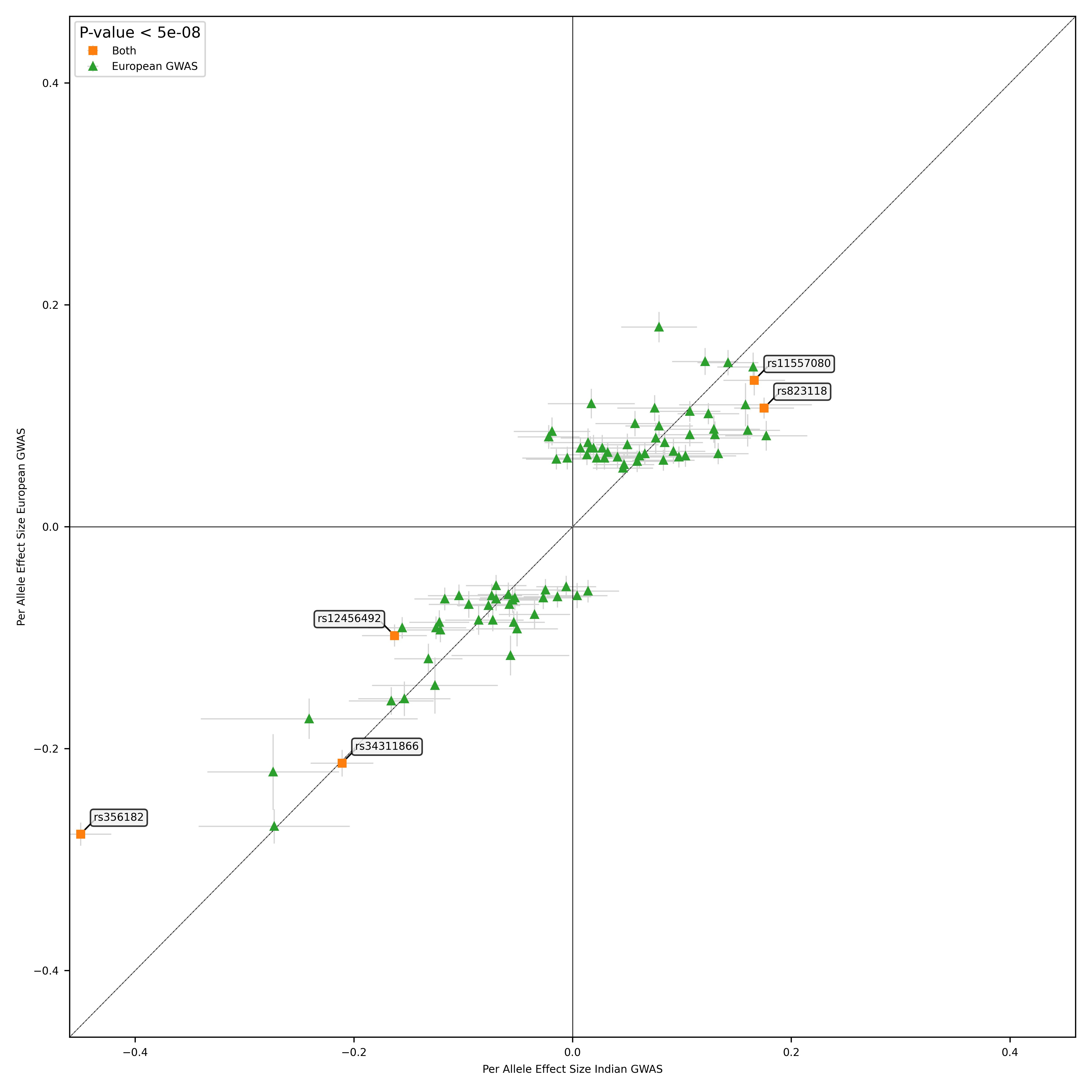

Effect estimates plot annotated with top SNPs from the European meta-analysis were plotted against the effect estimates from the corresponding SNPs effect estimates from the Indian GWAS. The X-axis indicates per-allele effect estimates of Indian GWAS. The Y-axis indicates per-allele effect estimates of the European meta-analysis.

**Supplementary Figure 15:** Manhattan plot highlighting signals from a meta-analysis of Indian GWAS and multi-ethnic PD meta-analysis. Three novel genes were identified.

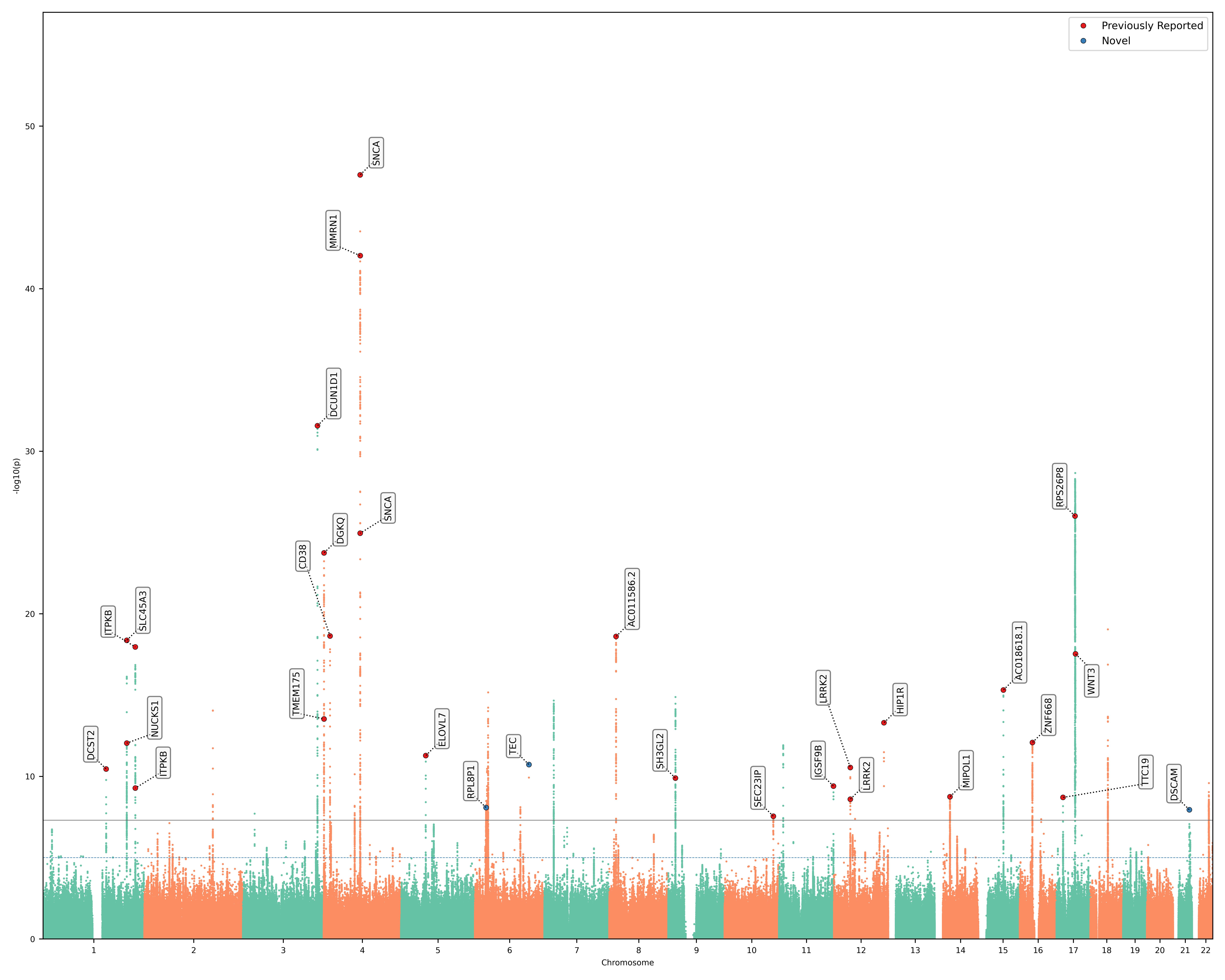

**Supplementary Figure 16:** MAGMA tissue expression analysis using the Indian-PD dataset on GTEx v8 53 tissue types

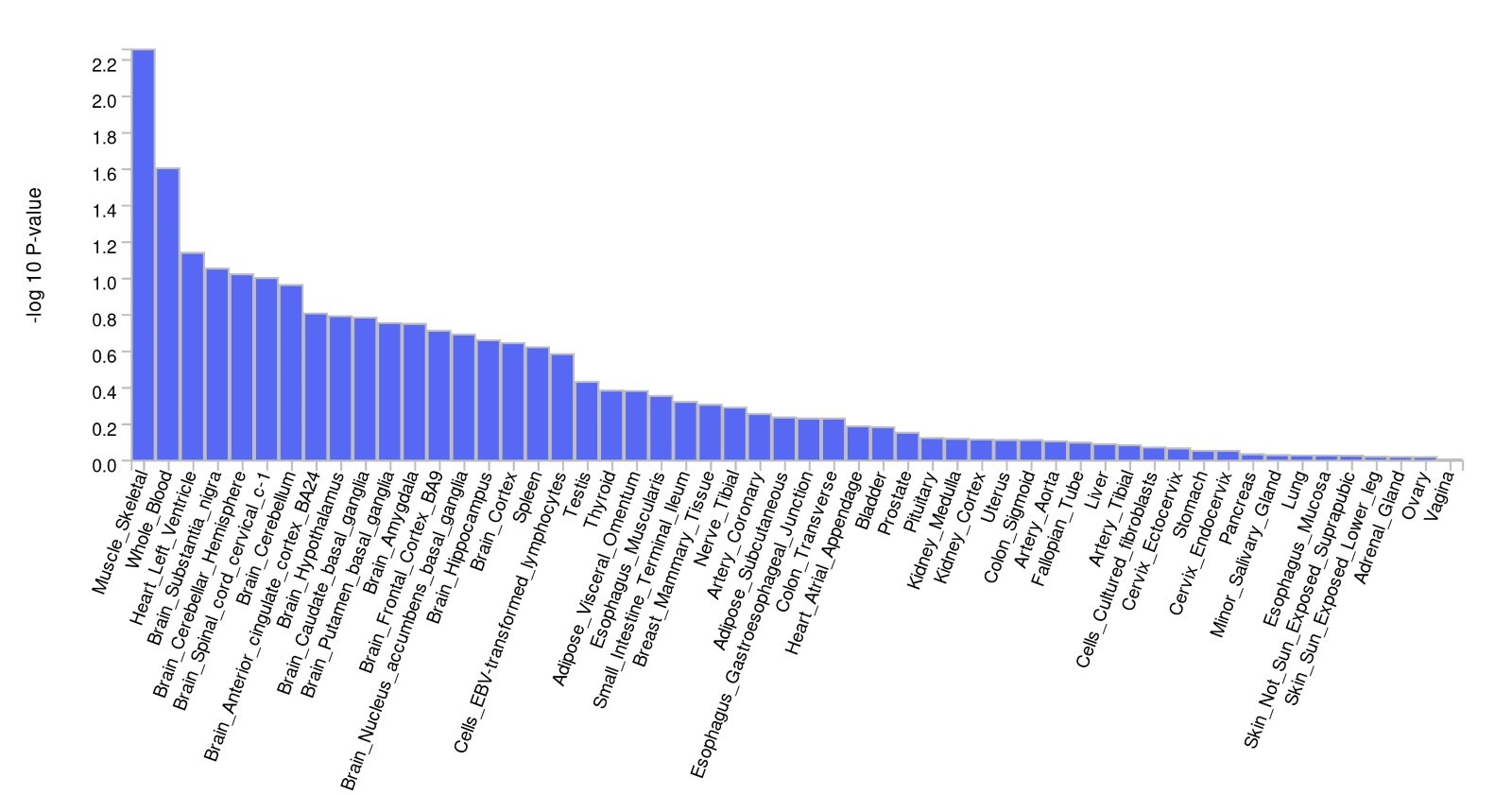

**Supplementary Figure 17:** Dedicated workflow and the corresponding organizational measures of IDEAL-GENOM pipeline to show the secure storage volumes to perform genomic analysis and provide access to the luxgiant resources to the scientific community.

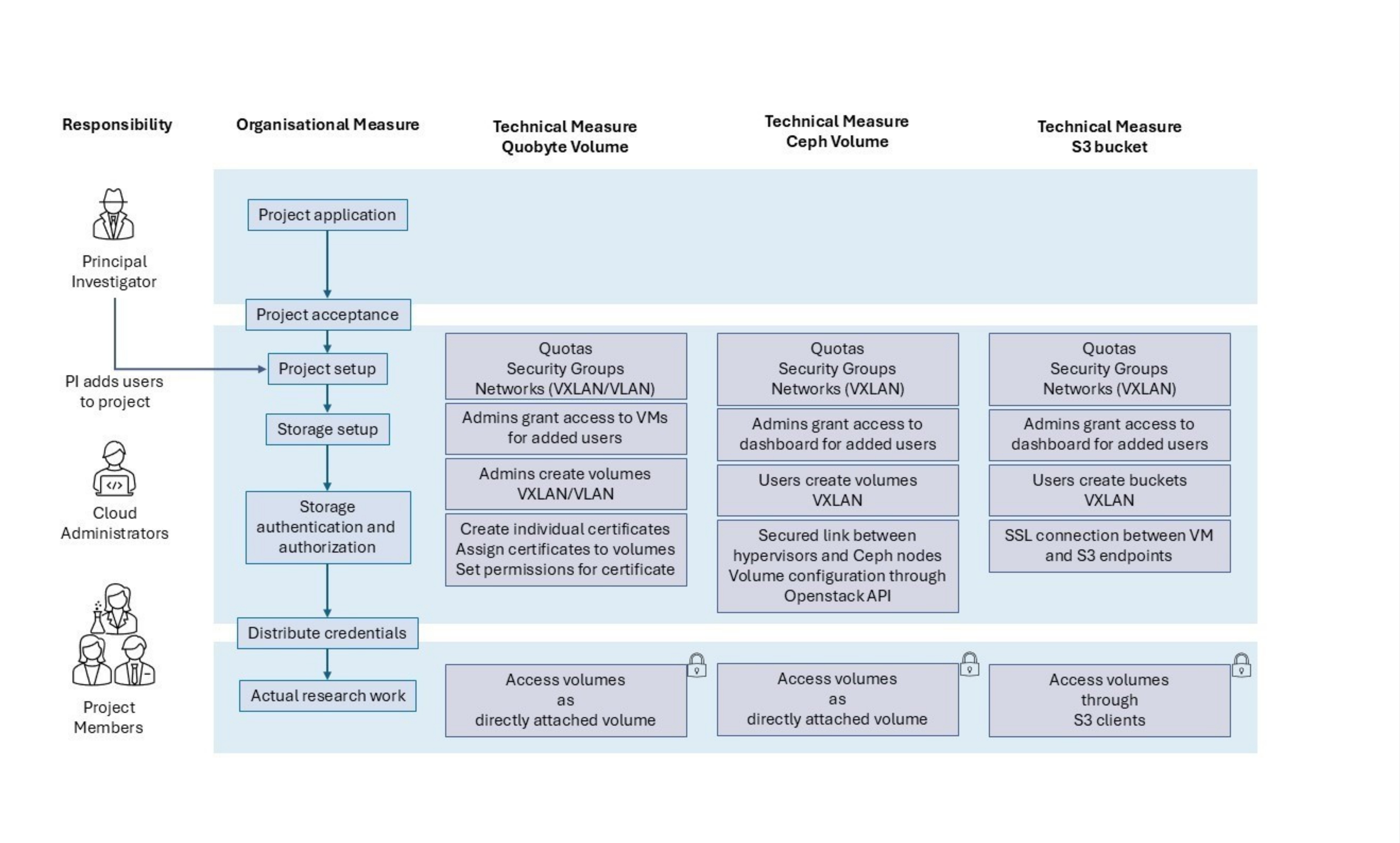
